# Efficacy of chloroquine and hydroxychloroquine in treating COVID-19 infection: a meta-review of systematic reviews and an updated meta-analysis

**DOI:** 10.1101/2020.07.28.20164012

**Authors:** Tawanda Chivese, Omran A. H. Musa, George Hindy, Noor Al-Wattary, Saif Badran, Nada Soliman, Ahmed T. M. Aboughalia, Joshua T. Matizanadzo, Mohamed M. Emara, Lukman Thalib, Suhail A. R. Doi

## Abstract

**Objective:** To synthesize findings from systematic reviews and meta-analyses on the efficacy and safety of chloroquine (CQ) and hydroxychloroquine (HCQ) with or without Azithromycin for treating COVID-19, and to update the evidence using a meta-analysis.

**Methods:** A comprehensive search was carried out in electronic databases for systematic reviews, meta-analyses and experimental studies which investigated the efficacy and safety of CQ, HCQ with or without Azithromycin to treat COVID-19. Findings from the reviews were synthesised using tables and forest plots and the quality effect model was used for the updated meta-analysis. The main outcomes were mortality, the need for intensive care services, disease exacerbation, viral clearance and occurrence of adverse events.

**Results:** Thirteen reviews with 40 primary studies were included. Two meta-analyses reported a high risk of mortality, with ORs of 2.2 and 3.0, and the two others found no association between HCQ and mortality. Findings from two meta-analyses showed that HCQ with Azithromycin increased the risk of mortality, with similar ORs of 2.5. The updated meta-analysis of experimental studies showed that the drugs were not effective in reducing mortality (RR 1.1, 95%CI 1.0-1.3, I^2^ =0.0%), need for intensive care services (OR 1.1, 95%CI 0.9-1.4, I^2^ =0.0%), virological cure (OR 1.5, 95%CI 0.5-4.4, I^2^ =39.6%) or disease exacerbation (OR 1.2, 95%CI 0.3-5.9, I^2^ =31.9%) but increased the odds of adverse events (OR 12,3, 95%CI 2.5-59.9, I^2^ =76.6%).

**Conclusion:** There is conclusive evidence that CQ and HCQ, with or without Azithromycin are not effective in treating COVID-19 or its exacerbation.

**Registration:** PROSPERO: CRD42020191353

## Introduction

The severe acute respiratory syndrome coronavirus 2 (SARS-CoV2) is currently known to be among one of the most contagious viruses in the history of pathogens (1). This novel human coronavirus was first reported in Wuhan in December 2019 and has since spread from person to person in an efficient and sustained way to cause a global pandemic, with the disease named Coronavirus Disease-19 (COVID-19) (1). Although the disease is largely controlled in China, where it was first reported, the number of cases and deaths continue to increase globally, with the highest numbers of confirmed cases in the USA, Brazil and India (2). The availability of vaccines has resulted in the relaxation of restrictive measures in many countries. However, the vaccinations have been slow in the low-and-middle income countries and the morbidity and mortality from COVID-19 are expected to continue increasing as successive waves driven by variants of the SARS-CoV-2 virus affect many countries (3-5). This continued increase in morbidity and mortality from COVID-19 warrants the need to investigate effective therapies.

Of several therapeutic drugs that have been suggested, chloroquine (CQ), an anti-malarial drug in the class of 4-aminoquinolones with anti-inflammatory, antiviral and anti-thrombolytic properties, and its derivative, hydroxychloroquine (HCQ) have been re-purposed and are being used widely for the treatment of COVID-19 (6, 7). CQ and HCQ are antimalarials and also used as disease-modifying antirheumatic drugs (DMARDs) (8). There are several mechanisms which have been suggested for the expected effect of the two drugs against viruses in general and SARS-COV-2 in particular. One of these mechanisms is based on inhibiting the ability of the virus to enter the cell. SARS-COV-2 harbours a spike (S) protein, which is considered a crucial element in the virus replication cycle as it binds to angiotensin-converting enzyme 2 (ACE2) expressed in the lungs of the host cell receptor (9). The antiviral effect of both CQ and HCQ is thought to be through their ability to interfere with the glycosylation of ACE2 and thus prevent the proper binding of the S protein (8). Moreover, virus entry occurs through receptor-mediated endocytosis, which needs an acidic PH to complete the fusion and deliver the viral genome into the cell. Both CQ and HCQ are weak bases and thought to inhibit this process that the SARS COV-2 virus needs for replication (8). Apart from their direct antiviral properties, CQ and HCQ also seem to have immune-modulatory effects which help to reduce over-activation of the immune system from COVID-19 (8). HCQ has been used more frequently than CQ in treating COVID-19 as it has been reported to be more potent in vitro (10), can be used in higher doses for a longer time with a lower risk of adverse events compared to CQ and is more widely available (6, 11). Further, both drugs are relatively cheap, and their use in the treatment of COVID19 is based on promising results from *in vitro* studies (10, 12) and some observational studies (13). Repurposing the drugs for use in treating COVID-19 has been easy as the drugs are already in use for the treatment of malaria, are cheap and have been thought to have a relatively safe profile (6, 14). Due to the above reasons and the unprecedented situation and the urgency to curb the COVID-19 pandemic, CQ and HCQ have been approved, on a fast track basis, by the United States Food and Drug Administration and other regulatory bodies, despite the lack of good quality evidence of their efficacy and safety (15, 16). Consequently, more than 80 trials of the two drugs are either ongoing or completed and it is necessary to evaluate the evidence so clinicians and regulatory bodies can make informed decisions on the use of these drugs for the treatment of COVID-19 infection.

Initial reports suggested that HCQ was effective in the treatment of COVID-19 associated pneumonia (17) and that HCQ in combination with the second-generation macrolide antibiotic, Azithromycin, resulted in a lower fatality (18), and 93% viral clearance of COVID-19, by day 8 (13). However, subsequent findings from both observational studies and clinical trials have been contradictory (19-22). Further, although the two drugs are relatively safe in the treatment of malaria, concerns have been raised about the risk of adverse events associated with their use when treating COVID-19 (23). The RECOVERY trial (24), one of the largest trials to-date investigating the optimal treatment for COVID-19, issued a statement that they found no differences in mortality between participants on HCQ and those on usual care, and subsequently stopped the HCQ arm of the trial. In a similar move, the World Health Organization stopped the HCQ arm of the SOLIDARITY trial (23), citing the lack of efficacy from their interim analysis of the U.K. RECOVERY trial, the French DisCoVeRy trial (25) and an unnamed Cochrane review (26). Before the stopping of the RECOVERY and SOLIDARITY trials, the largest observational study to date, a multinational cohort study of 96,000 participants, had previously reported a 6-to-8-fold increase in the mortality associated with CQ or HCQ, with or without macrolides (27). However, the study (27) was subsequently retracted due to concerns over the veracity of the data and its analysis, leaving some degree of uncertainty in its wake.

Several systematic reviews and meta-analyses investigating the efficacy and the safety of CQ and HCQ have been published (28-32). Similar to the primary studies, systematic reviews and meta-analyses have presented contradictory findings. A key issue that made it difficult to have conclusive results on the efficacy of CQ and HCQ with or without the macrolide antibiotics is that clinical trials were few, small and poorly designed. Most of the existing systematic reviews have therefore carried out meta-analyses combining data from observational and experimental studies without accounting for the quality of included studies. Several clinical trials, with acceptable quality, are now available. Therefore, it has become necessary to synthesize all available evidence to provide the best evidence-based assessment on the efficacy and safety of both CQ and HCQ, with or without macrolide antibiotics, in the treatment of COVID-19 infection. In this respect, we conducted this umbrella review with two broad aims; (1) to assess the efficacy and safety of each of HCQ and CQ, with or without Azithromycin in the treatment of COVID-19 by assessing the evidence from existing systematic reviews and meta-analyses, and (2) to carry out an updated meta-analysis of the existing experimental studies to assess the efficacy of these drugs.

## Methods

### Study Design

This study protocol is registered on PROSPERO (CRD42020191353). This study had two components; an overview of all existing systematic reviews and meta-analyses and an updated meta-analysis of all eligible experimental studies that investigated the efficacy of either CQ or HCQ with or without macrolide antibiotics in the treatment of COVID-19. The design of this overview followed the Preferred reporting items for overviews of systematic reviews including harms checklist guidelines (PRIO-harms) (33). The updated meta-analysis was carried out according to the Preferred Reporting Items for Systematic Review and Meta-Analysis (PRISMA) (34).

### Search strategy

We conducted electronic searches for all experimental studies, systematic reviews and meta-analyses on the efficacy and safety of CQ and HCQ for the treatment of COVID-19 up to the 3^rd^ of June 2020. We searched for studies in the following electronic databases; Cochrane Database of Systematic Reviews (CDSR) (*The Cochrane Library*), Cochrane Central Register of Controlled Trials (CENTRAL), the China Academic Journals Full Text Database, EMBASE and MEDLINE through PubMed, the Database of Abstracts of Reviews of Effectiveness (DARE), Scopus, Web of Science, CINAHL, and the databases of preprints. We hand-searched the databases of preprints (medrxiv.org/ and https://www.biorxiv.org/) and the websites for the World Health Organization Solidarity Trial and the U.K. Recovery Trial. We manually searched all references of all included studies and carried out a citation search of all included systematic reviews. We identified experimental studies from the included reviews and from the database searches. The top 20 similar articles citation search of all included studies was performed on Pubmed to retrieve studies that might have been missed in the original electronic search.

We used the following search terms for chloroquine and hydroxychloroquine; “chloroquine” OR “hydroxychloroquine” OR “CQ” OR “HCQ”. We used the following search terms for COVID19 infection; “COVID19” OR “Coronavirus” OR “novel coronavirus” OR “SARS-CoV-2” OR “COVID” OR “COVID-19”. The following search terms were used for the study design; “clinical study”, “clinical trial”, “trial”, “RCT”, “controlled trial”, “randomized controlled trial”, “meta-analysis”, “rapid review”, “review”, “systematic reviews”. All searches had no limitations on language or location but were limited to studies published during the year 2020. Keywords and Medical Subject Headings (Mesh) were used for searches for COVID19, SARS-CoV2, chloroquine and hydroxychloroquine. The full electronic search strategy is given in Appendix 1 (Supplementary Document S1).

### Eligibility

We included systematic reviews and meta-analyses which compared the efficacy and/or safety of HCQ or CQ. Such reviews should either have a minimum of two clinical studies comparing HCQ or CQ with or without Azithromycin to any other standard treatments, including placebo. Participants in the included index primary studies should have had confirmed COVID-19, regardless of age or the severity of illness. Reviews were excluded if they were literature reviews, did not include at least two eligible primary studies and if their main scope was on prophylaxis. The updated meta-analysis only included experimental studies which investigated the efficacy of either CQ, HCQ or without Azithromycin for the treatment of COVID-19. Studies where participants were diagnosed using symptoms were excluded as symptoms have been shown to have poor diagnostic accuracy (35). Observational studies and any studies without control groups were excluded. Studies on animals and in-vitro studies were also excluded. In the case that studies were duplicates, either the study with the most data was used and the other excluded or both studies were combined. Studies were included if the intervention included either CQ or HCQ alone or with Azithromycin in any dose combinations and any length of administration. Due to the heterogeneous nature of COVID-19 treatments in different countries and different disease severity categories, any studies that had control groups that did not include either CQ or HCQ were acceptable for inclusion in this overview and meta-analysis. Studies were also included irrespective of either the severity of disease of included participants or the setting (i.e. either hospital-based or community or both).

### Data extraction

Search results were uploaded on to the Rayyan systematic review management platform (https://www.rayyan.ai/) where 2 authors blindly screened titles and abstracts. Conflicts were discussed and resolved by consensus between authors. The full text of potentially relevant articles were screened against eligibility criteria for the final inclusion.

For each study, two authors independently extracted data and assessed quality. The data extracted from the reviews included; type of review (systematic review or meta-analysis), date of publication or submission to the preprint servers, countries with data included in the review, scope of the review, the number and type of index studies included, tools used to assess the risk of bias, risk of bias summary, main comparisons, total participants, mean age, outcomes measured and pooled measures of effect, statistics for heterogeneity, the review conclusion and limitations. Additional data included the number of and citations of all included primary studies in the review. The primary studies were further categorised as either experimental or observational, first based on their classification in the parent review and later after an independent assessment by two authors. All experimental studies identified from the assessment were considered for inclusion in the updated meta-analysis.

Data extracted from experimental studies included study design (RCT or quasi-RCT), intervention (whether CQ or HCQ with or without Azithromycin), dosage, route of administration, control treatment description, any co-interventions, setting (community setting or hospital), the country where study carried out, proportions with severe sickness, proportions with comorbidities, mean age, gender distribution, and total with each out outcome in the intervention and control groups.

### Outcomes

Due to the heterogeneous nature of the outcomes assessed in different systematic reviews, meta-analyses and primary studies, we grouped the outcomes into four main groups. The primary outcome was mortality assessed as all-cause mortality. Secondary outcomes were disease exacerbation, virological cure, adverse events and included a composite outcome that combined the events of any transfer to the intensive care unit (ICU), any need for intubation or any need for mechanical ventilation. Disease worsening was defined as any form of symptom worsening such as the need for oxygen, dyspnoea, hospitalization in the case of community-based studies. The virological cure was defined as a negative PCR any time after commencement of treatment. Safety was assessed as the occurrence of the known adverse events of CQ or HCQ or Azithromycin. These adverse events included gastric side effects such as diarrhoea and vomiting, ventricular tachycardia possibly because of QTc prolongation and headache, blurred vision and rash.

### Assessment of study quality

Two authors independently assessed the quality of each included review using the Assessing the Methodological Quality of Systematic Reviews (AMSTAR) tool (36). Each review had a maximum score of 11 if the methodological quality is good and zero if the methodological quality is poor. Any disagreements were resolved by discussion between the authors.

For the experimental studies, the MethodologicAl STandard for Epidemiological Research (MASTER) scale (37) was used to assess the methodological quality of studies across safeguards listed within 7 standards. The 7 standards assessed were; equal recruitment, equal retention, equal ascertainment, equal implementation, equal prognosis, sufficient analysis and temporal precedence (38). A kappa interrater agreement was calculated for each study and the quality safeguard counts averaged if the Kappa was at least 0.70. Where the Kappa was below 0.7, the authors resolved disagreements by discussion or by referring to a third assessor if they did not resolve. The quality counts were used to rank studies for inclusion in a quality adjusted meta-analysis.

### Synthesis of findings

Synthesis of findings from different reviews was done using a combination of a structured summary of findings from the reviews and presentation in forest plots (39). A table with the findings of each review for each outcome was presented. For outcomes were several meta-analyses were available, forest plots were used to show the magnitude of the effect, the 95% confidence intervals (95%CI) and the number of included studies. Where there were no available meta-analyses, findings from systematic reviews were compared narratively. The overall score from the AMSTAR quality assessment and the I^2^ heterogeneity score were incorporated in the interpretation of the findings of each review. The forest plots were created in R statistical software (40) AND A map showing the distribution of all the index studies included in the included reviews was created using Tableau software (41).

For the updated meta-analysis, the quality effects model (42) was employed to pool the estimates (odds ratios), as it is more robust and performs better than the random-effects model (43). In some studies where outcome assessed cases were zero, a post hoc continuity correction adding (0.5) to all cells was employed for valid estimation of the odds ratio and its variance. Forest plots were used to depict the results of the pooled analysis. The quality effects method uses a relative quality rank from each of the studies to modify the study’s variance weight, thereby incorporating the quality of the study quantitatively into the results. The results of random effects analyses were reported in the supplementary material for comparison only if there was heterogeneity as without this it defaults to a fixed effect model similar to the quality effects model. The Stata software program (44) was utilized for meta-analysis. Forest plots were used to present the pooled odds ratios and their confidence intervals (CIs) (45). The Cochran Q test p-value was used to test for and the I^2^ statistic to quantify heterogeneity (46). The I^2^ statistic measures consistency and is an indication of the variability in the estimates of the effects that are caused by heterogeneity instead of sampling error, and ranges from a 0% (no heterogeneity) to 100% (high heterogeneity). I^2^ values above 50% indicate substantial heterogeneity and above 75% high heterogeneity. Doi plots and the LFK index (47) were used for assessing publication bias as they are more easier to interpret than funnel plots and in this case funnel plots could not be used as they are not recommended when there are less than 10 studies in a meta-analysis (48).

### Ethics

Ethics approval was not required as the study used published data.

## Results

### Search results and characteristics of included reviews

The search identified 158 reviews and 111 were excluded by screening the title and abstract (Fig 1). Most reviews were excluded because they were either literature reviews or reviews which did not include completed clinical studies (Fig 1 and Supplementary Table S2). The remaining 47 reviews were screened for eligibility through reading the full text and 24 were provisionally included. One additional review was identified through manual searching of references and the citations search. The final included reviews were 13, of which 5 (38.5%) were systematic reviews and meta-analyses and the remainder systematic reviews only (Table 1).

**Fig.1:**
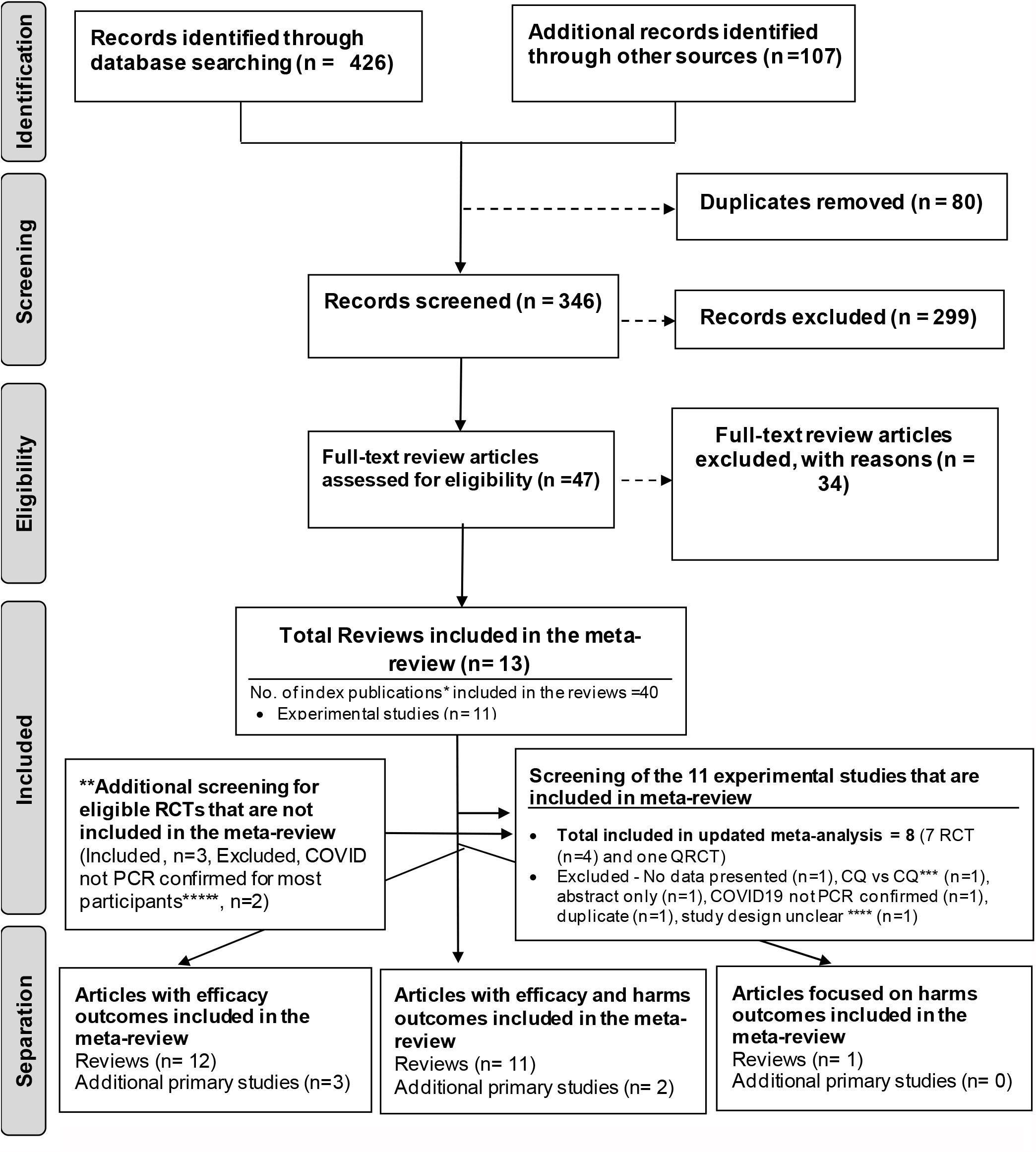
Flow chart for the meta-review. *Index publication is the first occurrence of a primary publication in the included reviews. **Additional eligible primary studies that had not been initially identified by the search of the relevant reviews or obtained by updating the search of the included reviews. ***study compared high dose chloroquine against a low dose chloroquine. **** study design not clear as both groups received HCQ, ***** most participants in study diagnosed using symptoms and not PCR

**Table 1.** Characteristics of included reviews.

| First author, date of publication/submission, study design | Scope of review | Types of included studies and total participants | Tool used to assess risk of bias | ROB summary |
| --- | --- | --- | --- | --- |
| Sarma (13 April 2020) (49)<br>Design - SRMA | Evaluation of safety and efficacy of HCQ alone or in combination | RCTs – 3<br>Observational – 4<br>Participants -1 358 | ROBINS-I tool, Newcastle Ottawa Scale | High risk of selection, performance and detection bias. Unclear risk of bias in attrition & reporting bias |
| Chowdhury (28 April 2020) (28)<br>Design - SR | To review the literature currently available regarding the clinical use of CQ and HCQ as treatment in COVID-19 | RCTs - 7<br>Total participants = 486 | Cochrane ROB tool | High risk (randomization, allocation concealment, and overall risk). 3 RCTs, 2 single arm, 1 observational |
| Singh (7 May 2020) (50)<br>Design - SRMA | Efficacy of HCQ in COVID-19 subjects on viral clearance and death due to all causes. | RCTs = 5 (2 non-randomized CT<br>Observational = 5<br>Total participants = 2042 | Jadad checklist, ROBINS I tool, Newcastle-Ottawa Scale | 5/8 on Jadad checklist, Moderate quality on ROBINS I tool (n=2), 7/8 on Newcastle-Ottawa Scale(n=2) |
| Suranagi (13 May 2020) (51)<br>Design - SR | To systematically explore, analyse, rate the existing evidence of hydroxychloroquine in the light of published, unpublished and clinical trial data. | RCTs- 3<br>Observational – 5<br>Total participants - 2047 | Oxford CEBM critical appraisal tool | Risk of bias was serious-very serious. Level of evidence quality/strength rating was GRADE level (low-very low) and CEBM level 2b-3b |
| Yang (14 May 2020) (52) | To demonstrate the significance of present | RCTs- 3 | The Modified Downs and Black risk | The average Downs and Black score were 19, with a |
| Design – SRMA | evidence regarding benefits and safety of HCQ for treatment of COVID-19 | Observational – 2<br>Total participants - 677 | assessment scale | range between 18 and 22 |
| Rodrigo (16 May 2020) (53)<br>Design – SR | To summarize evidence from human clinical studies for using HCQ or CQ as antiviral agents for any viral infection<br>20 agents for any viral infection | RCTs – 6<br>Total participants - 387 | Cochrane risk of bias tool and Robbins | Average score (3-5 out of 7).<br>Main types of bias were selection, reporting, performance and detection bias |
| Chacko (20 May 2020) (54)<br>Design – SRMA | To evaluate the efficacy and safety of hydroxychloroquine among patients with COVID-19 infection. | RCTs – 3<br>Observational – 8<br>Total participants 4306 | Cochrane risk of bias tool and Robbins-1 | Observational (3 low risk, 4 moderate, 1 high risk), RCT (risk of randomisation bias, allocation bias and blinding) |
| Hernandez (27 May 2020) (30)<br>Design – SR | Summarize evidence about the benefits and harms of HCQ or CQ treatment or prophylaxis of (COVID-19). | RCT – 3<br>Observational – 20<br>Total participants - 4186 | ROBINS-I and the Cochrane Risk of Bias 2.0 | Either no information or some concerns of bias to critical risk of bias |
| Shamshirian (28 May 2020) (55)<br>Design - SRMA | A review to overcome the controversies about the effectiveness of HCQ against COVID-19 | RCTs – 14<br>Observational – 7<br>Total participants - 103 486 | Jadad scale, ROBINS-I tool and Newcastle-Ottawa Scale | High risk of bias in randomisation sequence generation, allocation concealment, blinding and outcome assessment |
| Das (28 May 2020) (56)<br>Design – SR | Systematically review the therapeutic role of HCQ in COVID-19 | RCT – 4<br>Observational – 8 | NOS, Cochrane ROB tool | High risk of bias in most included studies |
| Takla (30 May 2020) (31)<br>Design – SR | Clarify the strength of evidence for the relative efficacy and safety of CQ and HCQ treatment | RCTs- 4<br>Observational – 15<br>Total participants - 107948 | Checklist of Review Criteria - Task Force of Academic Medicine and GERIME committee | All the studies included scored 13 in the risk assessment criteria. "12 were judged to be of scientific rigour" |
| Jankelson (31 May 2020) (23)<br>Design – SR | Review risk of QT prolongation, torsades, ventricular arrhythmia and sudden death with short courses of CQ and HCQ in COVID-19 | RCTs – 5<br>Observational – 6<br>Total participants - 1515 | None reported | Not reported |
| Wang (1 June 2020) (57)<br>Design – SR | Assess the published studies of Chloroquine (CQ) and hydroxychloroquine (HCQ) for the treatment of COVID-19 | RCTs – 10<br>Total participants - 1831 | Not reported | Not reported |

The characteristics of the included reviews and experimental studies are summarised in Table 1 and Table 2 respectively. The quality assessment of the included reviews and experimental studies are shown in Supplementary Tables 3 and 4, respectively.

**Table 2.** Characteristics of experimental studies included in the updated meta-analysis.

| Study and date of publication | Design and setting | Sample size and age (mean, years) | Drug and dose | Control | % with comorbidities | % of infection severity | Outcomes and length of follow-up |
| --- | --- | --- | --- | --- | --- | --- | --- |
| Huang et al., 18-May-2020 (58) | RCT, hospital<br>Country - China | 22 participants<br>Mean age 44.0 yrs (Range 36.5–57.5) | 10 participants received CQ (500 mg orally twice daily for 10 days) | 12 participants received Lopinavir/Ritonavir (400/100 mg orally twice daily for 10 days) | Not reported | 36.4% | -Negative conversion of SARS–CoV–2<br>- lung CT for assessing the improvement of NCP<br>-length of hospitalization<br>-Adverse events |
| Tang et al., 7-May-2020 (59) | Open-label RCT, 16 government-designed COVID-19 treatment centers<br>Country - China | 150 participants<br>Mean age 48.0 yrs (SD 14.1) | 75 participants received HCQ (1200mg for 3 days then maintained 800mg for rest of the follow up) + SOC | 75 participants received SOC only | 16% DM and 8% HTN | All participants had mild-moderate disease, except 2 were severe | -Negative conversion of SARS–CoV–2 by 28 days<br>-Adverse events |
| Chen Z et al., 10-April-2020 (60) | RCT (double-blind), Renmin Hospital of Wuhan University<br>Country - China | 62 participants<br>Mean age 44.7 (15.3) | 31 received HCQ (400 mg/d (200 mg/bid) between days 1 and 5) + SOC | 31 participants received SOC only | Not reported | 4 (6.5 %) had severe infection in control group | -Changes in TTCR<br>-Adverse events<br>-Mortality |
| Gautret et al., 20-March-2020 (20) | Open-label, quasi-experimental, 4 centres<br>Country - France | 42 participants<br>Mean age 45.1 (22.0) | 26 participants received HCQ (200mg, 3 times daily for 10 days)<br>6 participants received AZI (500mg day 1, followed by 250mg daily for the next 4 days) | 16 participants refused the protocol treatment | Not reported | 16.7% were asymptomatic, 61.1% with URTI and 22.2% with LRTI | -Viral clearance at day 6<br>-Mortality<br>-Adverse events |
| Chen J et al., 6-March-2020 (22) | Open-label RCT, Shanghai Public Health Clinical Center | 30 participants<br>Mean age 48.6 yrs (no SD reported) | 15 participants received oral HCQ (400mg once daily for 5 days) + SOC | 15 participants received SOC only | 36.7% with comorbid disease | Not reported | - Negative conversion of SARS–CoV–2 at day 7<br>-Disease exacerbation and adverse |

|  | Country – China |  |  |  |  |  | events |
| --- | --- | --- | --- | --- | --- | --- | --- |
| Horby et al. RECOVERY TRIAL 15- July 2020 (24) | Adaptive RCT<br>Country – U.K. | 4647 participants<br>Mean age 65.3 yrs (SD 15.3) | 1542 participants received a loading dose of HCQ of 800mg twice on the first day, then 400mg twice a day for 6 days | 3132 participants on SOC | 57% had at least one major comorbidity | 76% were either on mechanical ventilation or supplementary oxygen | Mortality after 28 days |
| Chen C.P. et al. 10 July 2020 (67) | RCT<br>Country – Taiwan | 33 participants<br>Mean age 32.9 yrs (SD 10.7) | 21 participants received 400mg HCQ twice a day on day one, then 200mg twice a day for 6 days | 12 participants on SOC | A “few” had comorbidities | Mild to moderate illness. Severe illness excluded. | Negative rRT-PCR at 14 days |
| Mitjà et al. 10 July 2020 (68) | RCT<br>Country - Spain | 293 participants<br>Mean age 41.6 yrs (SD 12.6) | 136 participants received 800mg on day 1, then 400mg once a day for 6 days | 157 participants on SOC | 53.2% | All had mild illness | -Reduction of viral load at 7 days<br>-Disease progression by day 28<br>-adverse events by day 28 |

*Abbreviations: n-RCT- nonrandomized clinical trial, q-RCT- quasi-randomized control trial, HCQ- hydroxychloroquine, SOC – Standard of care, DM- Diabetes mellitus, HTN, hypertension, CVD-cardiovascular diseases, CHF- chronic heart failure, CKF- chronic kidney failure, TTCR- time to clinical recovery, URTI-upper respiratory tract infection, LRTI-lower respiratory tract infection*

All the included reviews were conducted between 13 April and the 1^st^ of June 2020. At the time that this meta-review was carried out, 10 of the included reviews were published (23, 28, 30, 31, 49, 50, 52-55) and the remaining three (51, 54, 57)were preprints. All the included reviews assessed the efficacy and the safety of the CQ and HCQ in the treatment of COVID-19, except Jankelson et al. 2020 (23) which only assessed adverse events associated with CQ and HCQ.

Most of the included reviews had above 7 safeguards and only 2 reviews (23, 57) had 6 or lower safeguard counts on the AMSTAR scale (Supplementary Fig. 1 & Supplementary Table S3). The same 2 reviews did not report that they assessed for risk of bias in their included index studies. One (57) of the two reviews which did not report assessing the risk of bias in included studies was not published at the time of this umbrella review. The reported quality of included index studies in the reviews was generally poor, with 7 reviews (28, 49, 54, 56) reporting a low count of safeguards in general. The most common issues were lack of safeguards against selection bias, lack of randomization, lack of allocation concealment bias, absence of blinding and performance and reporting biases. Most of the included reviews had the limitations of a small number of included index studies, lack of proper control groups or improper randomization among the included primary studies. An additional limitation of the reviews was the mixing of results from observational and experimental studies in the meta-analyses of efficacy.

### Characteristics of primary studies included in reviews and the updated meta-analysis

The total number of index studies in the included reviews, after removing duplicate studies was 40. Most of the included primary studies were conducted in the USA (14 studies, 108 011 participants), followed by China (9 studies, 1975 participants) and France (9 studies, 1637 participants) (Fig 2). The total number of participants in all the reviews after removing duplicate studies was 113 786. All the included reviews included at least 3 experimental studies and the number of included observational studies ranged from zero in 3 reviews (28, 53, 57) to 20 in one review (55).

**Fig 2.**
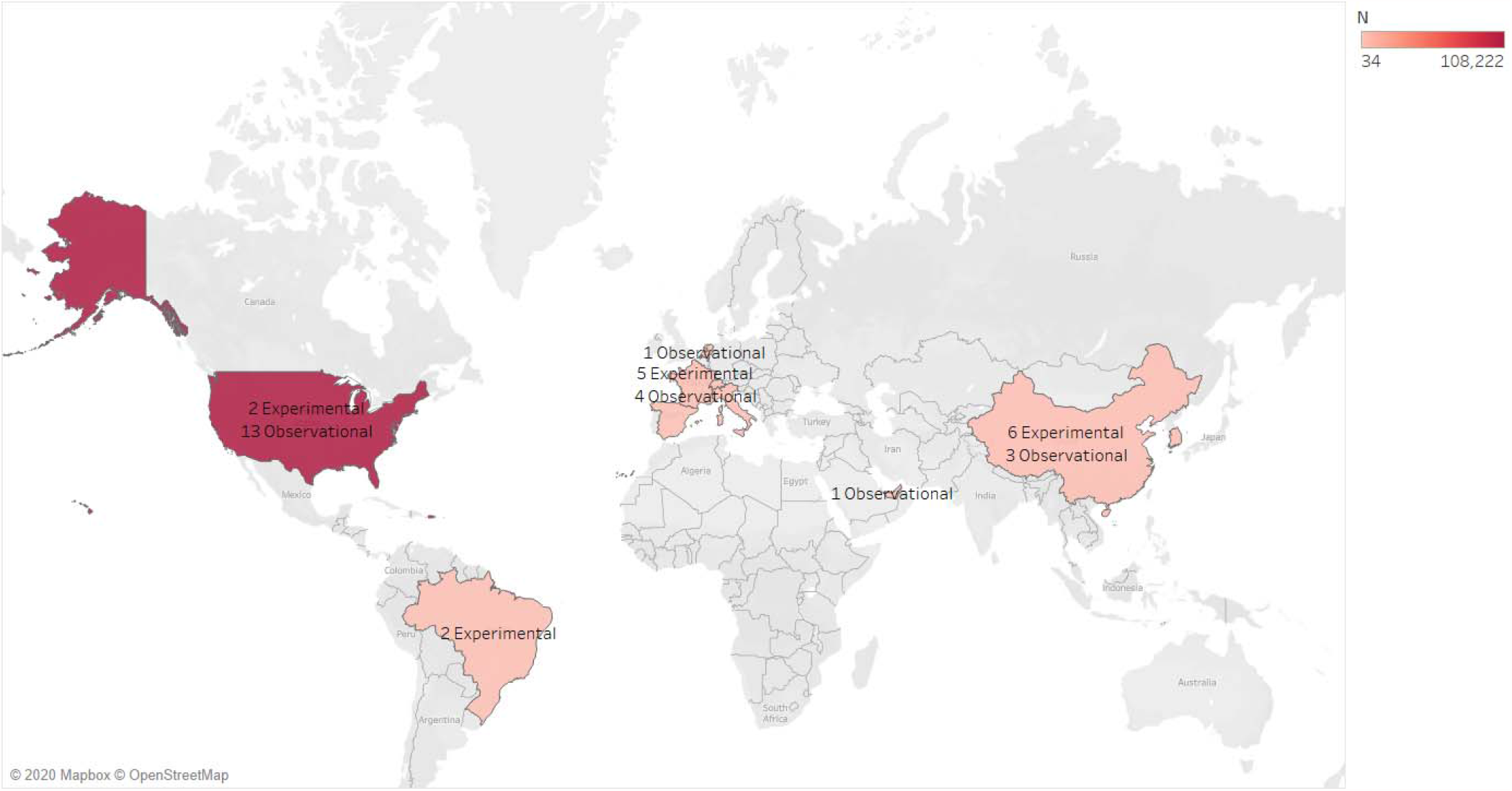
Location of all primary studies in included reviews. N – sample size of the index study

The observational index studies were excluded from the updated meta-analysis (Supplementary Table S4). After screening the 11 studies described as experimental studies in the included reviews, only 5 experimental studies (20, 58-60) satisfied the criteria for inclusion in the updated meta-analysis (Fig. 1). The reasons for exclusion of the other experimental studies were as follows; duplicate study (n=1) (61) which used data from Gautret et al. 2020 (20), one study had no full text available (62), one study compared high dose chloroquine to low dose chloroquine (63), and one study included participants who did not have COVID-19 confirmed by PCR (64). Another experimental study was excluded because it was a letter to the editor describing 15 trials with a total of 100 patients but with no clear details about the trials, the participants or any outcomes (65). Finally one study (66) was described by the authors as quasi experimental as two institutions treated patients confirmed as positive with HCQ but study subjects in both institutions received HCQ. One institution received HCQ shortly after admission, and in the other institution did not receive HCQ until days later when PCR results came through so there was no real control arm and was not published with a submission version of the pdf circulated on the internet. Five trials were identified from the additional search, with three included (24, 67, 68) and two (69, 70) excluded as they included participants who were diagnosed using symptoms rather than PCR (Fig. 1).

The eight experimental studies included in the updated meta-analysis (7 RCTs (22, 24, 58-60, 67, 68) and 1 quasi-experimental study (20)), had a total of 5279 participants and 1856 on either CQ/HCQ or HCQ with Azithromycin. Most of the studies (n=4) were from China (22, 58, 60, 67) and the remaining 4 studies each from France (n =1) (20), the U.K. (24), Spain (68) and Taiwan (59). Only one study used CQ (58) as the experimental drug while the remaining seven used HCQ. All seven (22, 24, 58-60, 67, 68) included RCTs had safeguard counts (quality assessment) of at least 29 out of 36. The quasi-experimental study (20) had the lowest safeguard counts of 16. All the included studies, except Chen Z et al. 2020 (60), either did not blind or did not report sufficient information about blinding of either participants and research performers.

Other notable deficiencies included insufficient reporting of randomisation procedure and insufficient statistical analyses (Supplementary Fig.1B & Supplementary Table S5).

### All-cause mortality

#### Findings from included reviews

A total of 4 meta-analyses, all with AMSTAR quality safeguard counts of at least 10, reported pooled effect sizes for mortality (49, 50, 54, 55) (Fig. 3). For HCQ alone, two meta-analyses; Yang et al. 2020 (52) (677 participants, 5 studies) and Singh et al. 2020 (50)(2042 participants, 10 studies) found a higher risk of mortality with HCQ with odds ratios of 3.0 and 2.2, respectively, with no heterogeneity (I^2^ = 0% for both). The other 2 reviews, Shamshirian et al. 2020 (55) (103 486 participants, 21 studies) and Charko et al. 2020 (54) (4306 participants, 11 studies) found slight but non-significant increases in the risk of mortality with HCQ use, with significant heterogeneity in both meta-analyses. Only 2 reviews (52, 55) reported pooled analyses on HCQ with Azithromycin and both found a higher risk of mortality associated with the use of HCQ with Azithromycin, with similar odds ratios of 2.5, with little heterogeneity in one and more heterogeneity in the other (Fig. 3).

**Fig 3.**
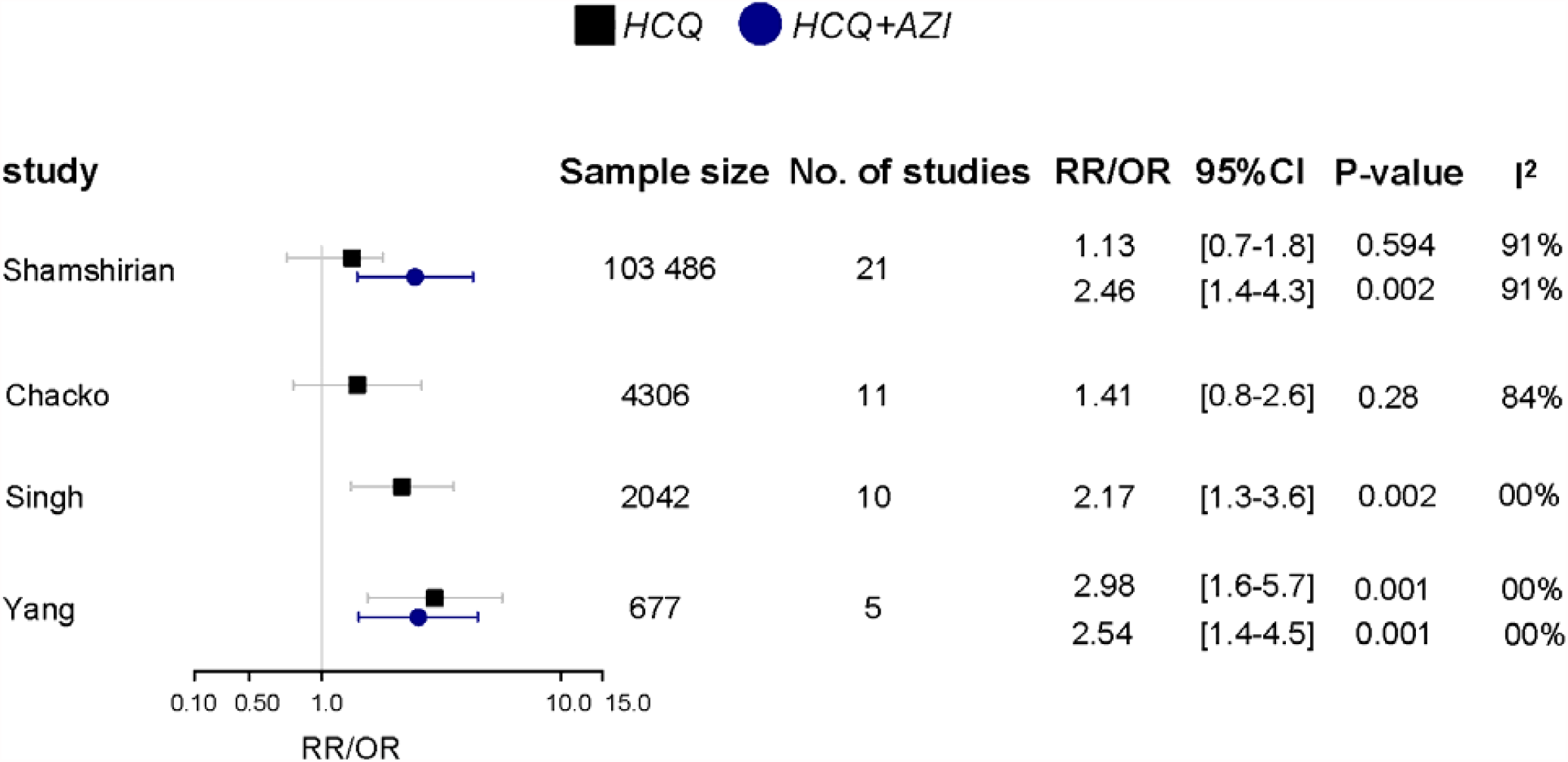
Results of meta-analyses - HCQ and all-cause mortality.

Of the reviews without meta-analysis, three concluded that there was evidence for higher risk of mortality in the HCQ group (30, 31, 51, 52) and one review (57) reported evidence of lower mortality in the HCQ group. Two reviews reported evidence of higher mortality in the chloroquine alone group (30, 53) (Supplementary Table S6).

#### Updated meta-analysis of experimental studies

Six experimental studies, five RCTs (22, 24, 59, 67, 68) and one quasi-experimental studies (20), assessed mortality, with a total of 5195 participants, of which 1815 were in the intervention group. There was no difference in the odds of mortality between participants who received HCQ with or without Azithromycin and those on standard care (OR 1.10, 95%CI 0.96 – 1.26; Fig 4), with consistency across studies (I^2^ = 0%).

**Fig. 4.**
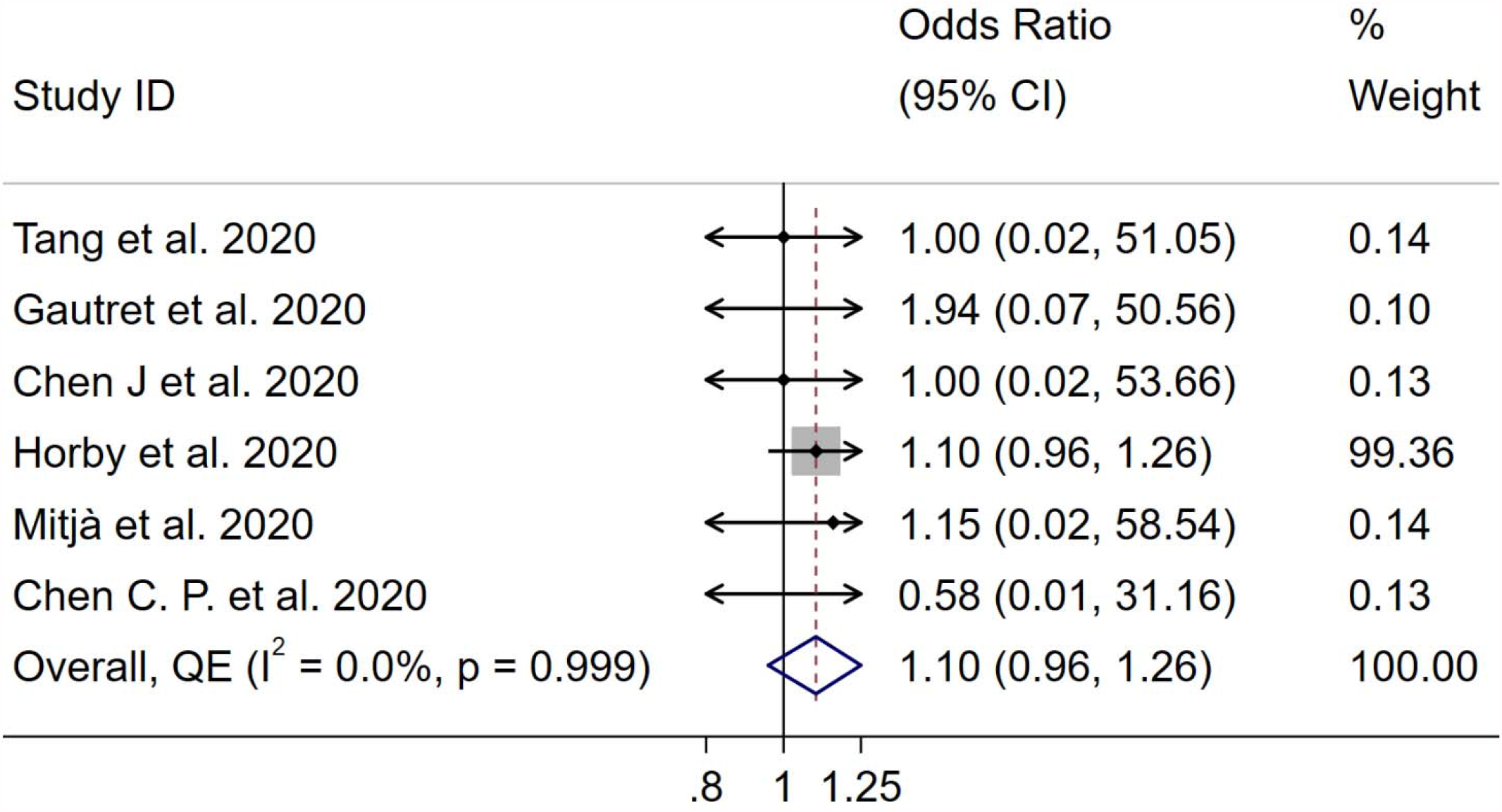
Updated meta-analysis of experimental studies – all-cause mortality. NOTE: Weights are from Doi’s Quality Effects model

The Doi plot (Supplementary Fig. 3A) showed minor asymmetry, indicative of possible publication bias. As Horby et al. 2020 (24) had the most weight in the meta-analysis, we carried out a sensitivity analysis without this study and the pooled odds ratio for the effect of HCQ with or without Axithromycin on all-cause mortality remained unchanged (OR 1.02, 95%CI 0.18 -5.86, I^2^ = 0.0%).

### Transfer to the ICU, intubation and mechanical ventilation

#### Findings from included reviews

A total of seven reviews investigated the risk of transfer to ICU, need for intubation and mechanical ventilation. Only one meta-analysis (55) of two studies with a total of 308 participants was done and found a non-significant 2-fold increase in the odds of intubation in individuals on HCQ (OR 2.11, 95%CI 0.31-14.03), with high heterogeneity between their included studies. The remaining six reviews (30, 31, 50, 51, 56, 57) which did narrative syntheses reported no effect of HCQ in the risk of transfer to the ICU, intubation or need for mechanical ventilation (Supplementary Table S6).

#### Updated meta-analysis of experimental studies

Two RCTs (24, 68), and the quasi experimental study (20), reported data on this outcome with a total of 4982 participants, of whom 1704 were in the intervention group. There was no significant difference in risk of ICU transfer, need for mechanical ventilation or intubation in participants who received HCQ with or without Azithromycin, compared to those on standard care (OR 1.11, 95%CI 0.88 – 1.41, I2 = 0.0%) (Fig. 5A). The Doi plot showed major asymmetry indicating possible publication bias (Supplementary Fig. 3B).

**Fig. 5.**
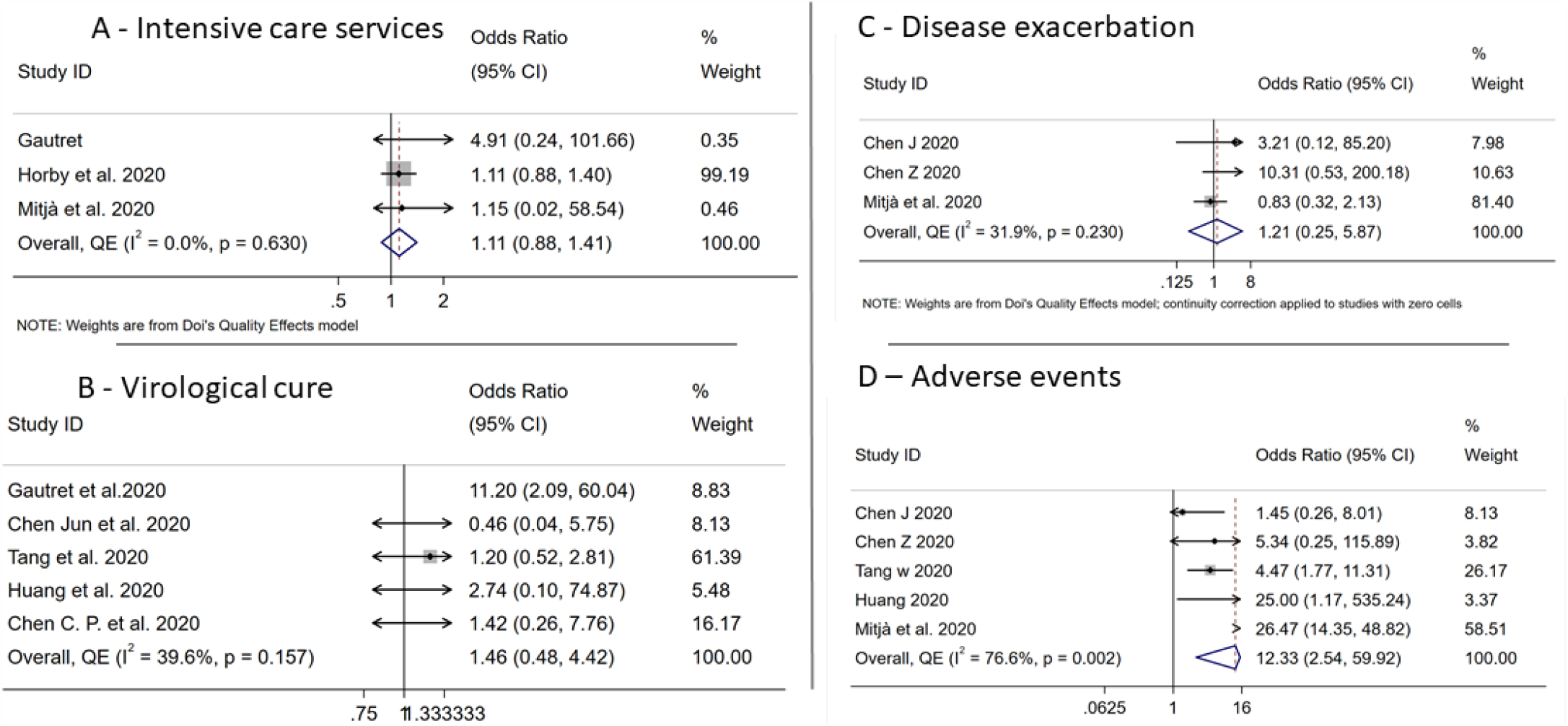
Updated meta-analysis of experimental studies - secondary outcomes.

### Virological cure

#### Findings from included reviews

Twelve reviews assessed the outcome of virological cure with 5 of the reviews being meta-analyses. All the 5 meta-analyses (49, 50, 52, 54, 55) found no differences between either HCQ alone or HCQ with Azithromycin and control, in virological cure (Fig. 6A).

**Fig. 6.**
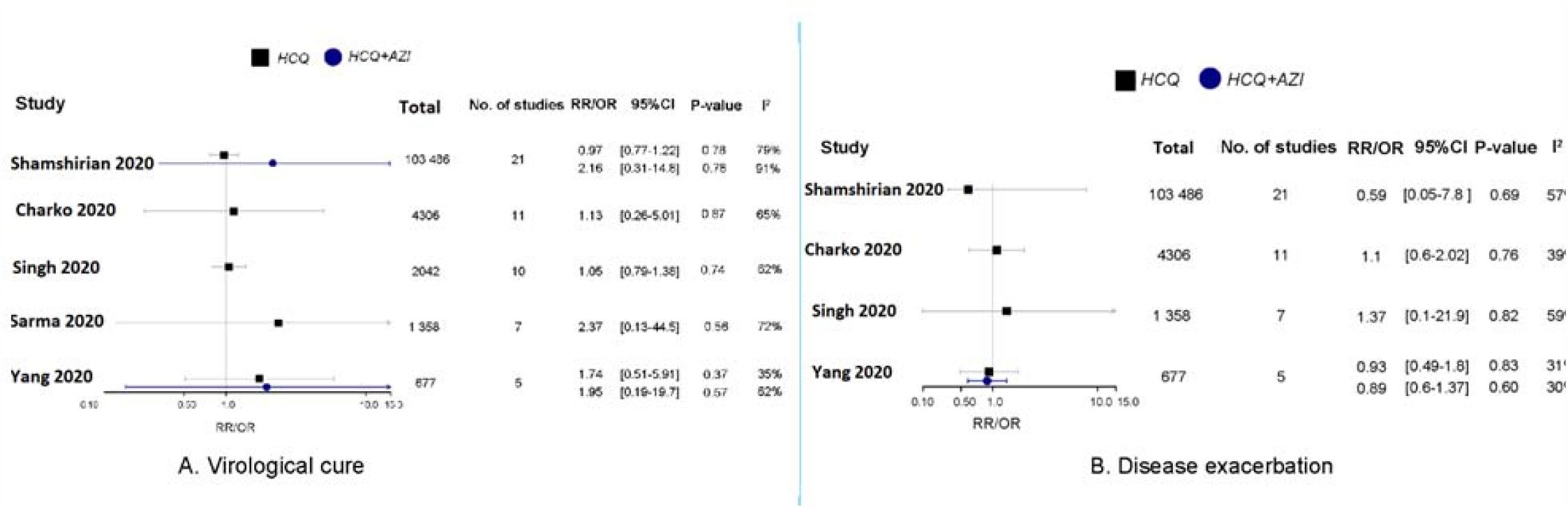
Results of meta-analyses on HCQ and virological cure and disease exacerbation.

However, the remaining seven reviews (28, 30, 31, 51, 53, 56, 57) which did narrative syntheses concluded that either HCQ alone or HCQ with Azithromycin were effective to some extent in the cure of the virus (Supplementary Table S6).

#### Updated meta-analysis of experimental studies

Five experimental studies, 4 RCTs (20, 22, 58, 59) and the single quasi-experimental study (20), assessed virological cure, with a total of 277 participants, of which 147 were in the intervention group. In pooled analyses, HCQ with or without Azithromycin appeared to improve virological cure although there was little evidence against the model hypothesis at this sample size (OR 1.46, 95%CI 0.48 – 4.48, I^2^ =39.6%) (Fig 5B). The Doi plot showed major asymmetry indicative of publication bias (Supplementary Fig. 3C). Removal of the quasi-experimental study, Gautret et al. 2020, did not alter the results of the pooled analysis (RR 1.02 95%CI 0.91-1.14, I^2^ =0%).

### Disease exacerbation

#### Findings from reviews

Four meta-analyses (50, 52, 54, 55), all with AMSTAR scores above 9, found that HCQ with or without Azithromycin had no effect on disease exacerbation (Fig 6B). The remaining reviews without meta-analysis on this outcome concluded that either HCQ or HCQ with Azithromycin reduced the severity of illness (Supplementary Table S6). Three reviews (50, 51, 57) concluded that pneumonia was improved in the HCQ with or without Azithromycin arm.

#### Updated meta-analysis of experimental studies

Three RCTs (22, 60, 68) assessed disease exacerbation, with a total of 385 participants, of which 182 were in the intervention group. The pooled analyses showed that HCQ with or without Azithromycin had no effect on disease exacerbation (OR 1.21, 95%CI 0.25 – 5.87, I^2^ =31.9%) (Fig. 5C). The Doi plot showed major asymmetry.

### Adverse events

#### Findings from reviews

A total of ten reviews investigated the risk of adverse events between HCQ or HCQ with Azithromycin groups and control. Two of these carried out meta-analyses and found pooled Odds Ratios of 3.9 (95%CI 1.8-8.1, n= 4 studies, 304 participants, I^2^ = 7.7%) (55) and 4.1 (95%CI 1.4-11.9, n= 4 studies, 1714 participants, I^2^ = 81%) (54). All the remaining 8 reviews (23, 28, 31, 49, 51, 56, 57) except one (30), found an increased risk of adverse events in the HCQ with or without Azithromycin group. The most reported adverse events were QTc interval prolongation diarrhea, arrythmia and first-degree AV block (Supplementary Table S6).

#### Updated meta-analysis of experimental studies

Five RCTs (22, 58-60, 68) assessed adverse events, with a total of 557 participants, of which 262 were in the intervention group. The most commonly reported adverse events due to HCQ and CQ were gastrointestinal (nausea, vomiting, diarrhea and abdominal pain), reported in all five trials (22, 58-60, 68), headache (60, 67, 68) and itchiness and rash (58, 60). Serious adverse events were very rare and reported in only three participants in three of the included trials (22, 24, 59). These included a single case of torsades de pointes reported in the RECOVERY trial (24), two cases, one with blurred vision and the other with thirst, reported by Tang et al. 2020 (59) and a case described as “severe” but with no clear details, Chen J et al. 2020 (22). The pooled odds ratio showed a 12-fold increase in the odds of adverse events in participants who received HCQ with or without Azithromycin, compared to those on standard care (OR 12.33, 95%CI 2.54 – 59.92, I2 = 76.6%) (Fig. 5D). The Doi plot showed major asymmetry indicative of possible publication bias (Supplementary Fig. 3E).

#### Conclusions from reviews

Nine of the reviews (28, 30, 50, 51, 53-57) all concluded that there was insufficient evidence to support the use of either CQ or HCQ with or without Azithromycin in the treatment of people with COVID-19. Two reviews concluded that there was some benefit in using HCQ with or without Azithromycin. Yang et al. 2020 (52) concluded that, although they were associated with higher mortality, HCQ with or without Azithromycin were beneficial, based on their effect on COVID-19 viral clearance. Sarma et al. 2020 (49) concluded that HCQ was beneficial based on its efficacy in reducing radiological progression and that the drug was safe. The remaining 2 reviews concluded that either CQ or HCQ were unsafe based on higher risk of mortality (31) and adverse events (23, 31).

## Discussion

In this meta-review, we summarized the findings of seven systematic reviews and five meta-analyses and carried out an updated meta-analysis to investigate the efficacy and safety of CQ and HCQ, with or without a second-generation macrolide antibiotic, Azithromycin, in individuals infected with COVID-19, limiting our analysis to eight experimental studies which met a strict inclusion criterion. Findings from the included reviews suggested that HCQ or CQ with or without Azithromycin had no benefit; two studies suggesting a 2-to-3-fold increase in the risk of mortality in those taking the drugs, while the other two reviews found no association with mortality. Only two meta-analyses were carried out for HCQ combined with Azithromycin and both reported a 2.5-fold increase in risk of mortality. The updated meta-analyses carried out for this review showed that HCQ, with or without Azithromycin, was not beneficial in reducing the risk of mortality from COVID-19. Findings from included reviews suggested no beneficial effect of either HCQ alone or HCQ with Azithromycin on the need for intensive care services, disease exacerbation and virological cure. Our updated meta-analyses also showed that there was no beneficial effect of HCQ with or without Azithromycin on the risk of transfer to the ICU, intubation or need for mechanical ventilation, virological cure and disease exacerbation. Lastly, all the included reviews, except one, found that HCQ with or without Azithromycin increased the risk of adverse events, in agreement with our updated meta-analysis.

An important consideration in this meta-review is the impact of methodological limitations on the results of both the primary studies and the systematic reviews and meta-analyses included in this umbrella review. These limitations could primarily have resulted from the urgency of the need to find a cure, at short notice, for a pandemic that seemed to be worsening in many countries. The limitations include, but are not limited to, small study sample sizes, the scarcity of randomized controlled trials, and the lack of methodological rigueur in the primary studies. All the reviews, except one (28), included observational studies, which tend to have confounding and may lead to biased estimates of effects. An additional weakness of these observational studies is that patients and clinicians will most likely choose an experimental drug, compared to standard care which comprises of symptom management, during a pandemic with a perceived high risk of death and no cure. Faced with a life-threatening illness, patients with severe illness will likely choose the experimental drug in the absence of proven alternatives, while those with mild to moderate disease may not want the experimental drug. The inclusion of observational studies in these meta-analyses seems to have been driven by the lack of good quality experimental studies and the need to use as much of the available information as possible. The observational studies were bigger than the RCTs and therefore tended to influence the pooled estimates. The effect of inclusion of the observational studies could have been reduced by using quality-adjusted meta-analysis synthesis, which decreases the weight of the observational studies in the overall estimate. However, none of the existing meta-analyses adjusted for the quality of the included studies in their syntheses, except Shamshirian et al. 2020 (55), who carried out a sensitivity analysis with experimental studies only. Further, the poor quality of studies included in the reviews is one of the limitations frequently cited by the review authors. Some major limitations in the experimental studies include the inclusion of participants who had COVID-19 diagnosed using symptoms in one study (64), controlled clinical trials which had a high risk of selection bias (66), and studies where the veracity of data presented could not be verified (71). An example of the later is a letter about 15 clinical trials in China claiming that CQ was effective (27, 71) but without any data presented. This letter was included in two reviews (54, 57) and contributed to the perceived efficacy of CQ and HCQ, which resulted in regulatory approvals in many countries. The risk of bias associated with a lack of either proper randomization or protection of the allocation sequence in the controlled trials is particularly serious in the case of COVID-19. This is because without an effective cure, and based on the hype about CQ/HCQ, severely ill patients were more likely to be given the experimental treatment, and consequently have worse outcomes if the treatment was not efficacious. Finally, it would be remiss to not mention that the biggest observational study included in two of the meta-analyses (31, 55) was subsequently retracted (27). This impacts the findings of these reviews although one of the reviews carried out a sensitivity analysis without this study.

The findings of our updated meta-analysis of experimental studies showed no benefit of HCQ, with or without Azithromycin in reducing the risk of all-cause mortality. Our findings are in agreement with findings of two of the included meta-analyses (54, 55) while two other reviews (50, 52) found a higher risk of mortality in the HCQ arms. Of the two meta-analyses which found a higher risk of mortality, Singh et al. 2020 (50) included three studies with 474 participants from two observational studies (72, 73) and the one quasi-experimental study (66) had a weight of 4.6%, in their meta-analysis. Yang et al. 2020 (52) included data from two small trials and one observational study, the US veterans study (72) which had a weight of 95% in the meta-analysis, and is misclassified as an RCT in the meta-analysis. Further, all the deaths, except one, occurred in the U.S. veterans’ observational study, and therefore the meta-analysis effectively drew its conclusions from the one big observational study where clinical severity may have influenced allocation of treatment. Similar results were also observed by Yang et al. 2020 (52) who also included the same three studies in their meta-analysis. Although Shamshirian et al. 2020 (55) found no significant difference in mortality when HCQ was used alone, they found a 2.5 fold increase in risk of mortality when the drug was combined with Azithromycin, a result similar to that of Yang et al. 2020 (52). The meta-analysis by Shamshirian et al. 2020 included several observational studies and data from the retracted study by Mehra et al. 2020, although sensitivity analyses without this study did not alter their conclusions. Charko et al. 2020 (54) included data from five observational studies and only one quasi experimental study which had a weight of 5.8% in the meta-analysis. Despite these shortcomings in the reviews, the findings from our updated meta-analysis and these existing reviews suggest that, at the very least, HCQ with or without Azithromycin do not have a protective effect against mortality in individuals with COVID-19 and may be harmful, in the worst-case scenario.

The findings of this review also shows that HCQ with or without Azithromycin does not have a beneficial effect on other clinical outcomes. Our updated meta-analyses, in agreement with most of the included reviews (30, 31, 50, 51, 55, 56), showed no benefit of HCQ with or without Azithromycin in reducing the need for intensive care services, limiting disease exacerbation or viral clearance. Lastly, in the updated meta-analysis, and in agreement with most of the included reviews, HCQ with or without Azithromycin, was associated 12-fold increase in the odds of adverse events. It should be noted however, that the occurrence of serious adverse events in the included experimental studies was rare, in agreement with the known safety profile of both CQ and HCQ.

Our updated meta-analysis had limitations which include small sample sizes in seven of the eight included trials and high risk of selection bias in the included quasi-experimental study. Some of the strengths of this updated meta-analysis include the inclusion of data of individuals with confirmed COVID-19 only, the inclusion of experimental studies only and the use of quality effects models to adjust for the weight of the studies in the meta-analysis.

## Conclusion

The use of HCQ with or without Azithromycin, for treating COVID-19, does not seem to have benefit in reducing mortality or the severe sequela of COVID-19, including transfer to the ICU, intubation, mechanical ventilation, virological cure or disease exacerbation. Rather use of these drugs is associated with a higher risk of adverse events, mainly gastrointestinal such as vomiting, diarrhea, and nausea. These findings do not support any further use of either CQ or HCQ, with or without Azithromycin, for the treatment of COVID-19.

## Supporting information

Supplementary Document S1. Search Strategy

## Data Availability

Data are available upon request

## Author contributions

TC – Conception and design of study, data curation (search for studies, data extraction and assessment of quality) formal analysis, software, writing original draft and revisions and final approval of submitted draft

OAHM - Data curation (search for studies, data extraction and assessment of quality), formal analysis, software, revisions and final approval of submitted draft

GH - Revisions and final approval of submitted draft

NW - Data curation (data extraction and assessment of quality), revisions and final approval of submitted draft

SB - Revisions and final approval of submitted draft

NS - Data curation (data extraction and assessment of quality), formal analysis, software, revisions and final approval of submitted draft

ATMA - Data curation (data extraction and assessment of quality), formal analysis, software, revisions and final approval of submitted draft

JTM - Data curation (search for studies), revisions and final approval of submitted draft MME - Revisions and final approval of submitted draft

LT - Conception and design of study, formal analysis, revisions and final approval of submitted draft

SARD - Conception and design of study, data curation, formal analysis, software, revisions and final approval of submitted draft

## Abbreviations

COVID-19: Coronavirus Disease-19
SARS-CoV-2: severe acute respiratory syndrome coronavirus 2
HCQ: hydroxychloroquine,
CQ: chloroquine
PRIO-harms: Preferred Reporting Items for Overviews of Systematic Reviews including harms checklist
AMSTAR: Assessing the Methodological Quality of Systematic Reviews
MASTER: MethodologicAl STandard for Epidemiological Research
PRISMA: Preferred Reporting Items for Systematic Review and Meta-Analysis
CDSR: Cochrane Database of Systematic Reviews
CENTRAL: Cochrane Central Register of Controlled Trials
DARE: Database of Abstracts of Reviews of Effectiveness

