## Supplementary Document S1. Search Strategy for "Efficacy of chloroquine and hydroxychloroquine in treating COVID-19 infection: a meta-review of systematic reviews and an updated meta-analysis"

### Supplementary Information

#### Supplementary Document S1. Search Strategy

##### PubMed Search

A. Search 1 HCQ

1. HCQ OR Hydroxychloroquine OR + chloroquine OR CQ

B.Search COVID-19

2. COVID19 OR Coronavirus OR novel coronavirus OR SARS-CoV-2 OR COVID OR COVID-19

C.Combine 2 AND 3

C.Apply filters

-January 2020 – May 30 2020

- Systematic reviews, reviews, meta-analysis, clinical trial, randomized controlled trials and trials

Search: **HCQ OR Hydroxychloroquine OR chloroquine OR CQ** Filters: **Clinical Study, Meta-Analysis, Randomized Controlled Trial, Review, Systematic Reviews, in the last 1 year**
"HCQ"[All Fields] OR "hydroxychloroquine"[MeSH Terms] OR "hydroxychloroquine"[All Fields] OR "chloroquin"[All Fields] OR "chloroquine"[MeSH Terms] OR "chloroquine"[All Fields] OR "chloroquine s"[All Fields] OR "chloroquines"[All Fields] OR "crit q"[Journal] OR "cost qual"[Journal] OR "cost qual q j"[Journal] OR "commun q"[Journal] OR "caribb q"[Journal] OR "camb q healthc ethics"[Journal] OR "cq"[All Fields]

**Translations**

**Hydroxychloroquine:** "hydroxychloroquine"[MeSH Terms] OR "hydroxychloroquine"[All Fields]

**chloroquine:** "chloroquin"[All Fields] OR "chloroquine"[MeSH Terms] OR "chloroquine"[All Fields] OR "chloroquine's"[All Fields] OR "chloroquines"[All Fields]

**CQ:** "Crit Q"[Journal:__jid101653381] OR "Cost Qual"[Journal:__jid101126987] OR "Cost Qual Q J"[Journal:__jid9602863] OR "Commun Q"[Journal:__jid101580483] OR

"Caribb Q"[Journal:__jid101553695] OR "Camb Q Healthc Ethics"[Journal:__jid9208482] OR "cq"[All Fields]

Search: **COVID19 OR Coronavirus OR novel coronavirus OR SARS-CoV-2 OR COVID OR COVID-19** Filters: **Clinical Study, Meta-Analysis, Randomized Controlled Trial, Review, Systematic Reviews, in the last 1 year**

((((("covid 19"[Supplementary Concept] OR "covid 19"[All Fields]) OR "covid19"[All Fields]) OR (("coronavirus"[MeSH Terms] OR "coronavirus"[All Fields]) OR "coronaviruses"[All Fields])) OR ((("novel"[All Fields] OR "novel s"[All Fields]) OR "novels"[All Fields]) AND (("coronavirus"[MeSH Terms] OR "coronavirus"[All Fields]) OR "coronaviruses"[All Fields]))) OR (("severe acute respiratory syndrome coronavirus 2"[Supplementary Concept] OR "severe acute respiratory syndrome coronavirus 2"[All Fields]) OR "sars cov 2"[All Fields])) OR "COVID"[All Fields]) OR ((((((("covid 19"[All Fields] OR "covid 2019"[All Fields]) OR "severe acute respiratory syndrome coronavirus 2"[Supplementary Concept]) OR "severe acute respiratory syndrome coronavirus 2"[All Fields]) OR "2019 ncov"[All Fields]) OR "sars cov 2"[All Fields]) OR "2019ncov"[All Fields]) OR (("wuhan"[All Fields] AND ("coronavirus"[MeSH Terms] OR "coronavirus"[All Fields])) AND (2019/12/1:2019/12/31[Date - Publication] OR 2020/1/1:2020/12/31[Date - Publication])))

**Translations**

**COVID19:** "COVID-19"[Supplementary Concept] OR "COVID-19"[All Fields] OR "covid19"[All Fields]

**Coronavirus:** "coronavirus"[MeSH Terms] OR "coronavirus"[All Fields] OR "coronaviruses"[All Fields]

**novel:** "novel"[All Fields] OR "novel's"[All Fields] OR "novels"[All Fields]

**coronavirus:** "coronavirus"[MeSH Terms] OR "coronavirus"[All Fields] OR "coronaviruses"[All Fields]

**SARS-CoV-2:** "severe acute respiratory syndrome coronavirus 2"[Supplementary Concept] OR "severe acute respiratory syndrome coronavirus 2"[All Fields] OR "sars cov 2"[All Fields]

**COVID-19:** "COVID-19"[All Fields] OR "COVID-2019"[All Fields] OR "severe acute respiratory syndrome coronavirus 2"[Supplementary Concept] OR "severe acute respiratory syndrome coronavirus 2"[All Fields] OR "2019-nCoV"[All Fields] OR

"SARS-CoV-2"[All Fields] OR "2019nCoV"[All Fields] OR (("Wuhan"[All Fields] AND ("coronavirus"[MeSH Terms] OR "coronavirus"[All Fields])) AND (2019/12[PDAT] OR 2020[PDAT]))

#### Scopus

TITLE-ABS-KEY(1. HCQ OR Hydroxychloroquine OR chloroquine OR CQ) AND (COVID19 OR Coronavirus OR novel coronavirus OR SARS-CoV-2 OR COVID OR COVID-19) AND (REVIEW OR SYSTEMATIC REVIEW OR META-ANALYSIS OR METAANALYSIS OR METAANALYSES OR RCT OR CLINICAL STUDY OR RANDOMIZED CONTROLLED TRIAL OR CLINICAL TRIAL) AND ( LIMIT-TO ( PUBYEAR,2020) )

#### CINAHL

(HCQ OR Hydroxychloroquine OR chloroquine OR CQ) AND (COVID19 OR Coronavirus OR novel coronavirus OR SARS-CoV-2 OR COVID OR COVID-19) AND (REVIEW OR SYSTEMATIC REVIEW OR META-ANALYSIS OR METAANALYSIS OR METAANALYSES OR RCT OR CLINICAL STUDY OR RANDOMIZED CONTROLLED TRIAL OR CLINICAL TRIAL) AND ( LIMIT-TO ( PUBYEAR,2020) )

**Supplementary Table S2. Characteristics of excluded reviews**

| Study and year | Date of publications | Title | Objective | Design | Reason of exclusion |
|---|---|---|---|---|---|
| Cortegiani et al., 2020 | 10-March-2020 | A systematic review on the efficacy and safety of chloroquine for the treatment of COVID-19 | To summarize the evidence regarding chloroquine for the treatment of COVID-19. | Systematic review (SR) | Did not include completed clinical studies on COVID-19 |
| Zhu et al., 2020 | 17-March-2020 | Systematic Review of the Registered Clinical Trials of Coronavirus | To analyze the characteristics and existing problems of the registered clinical trials | Systematic review (SR) | Did not include completed clinical studies on COVID-19 |
| Singh et al., 2020 | 22-March-2020 | Chloroquine and hydroxychloroquine in the treatment of COVID-19 with or without | The efficacy of chloroquine and hydroxychloroquine, in the | Narrative review | Did not include completed |

| | | | | | |
|---|---|---|---|---|---|
| | | diabetes: A systematic search and a narrative review with a special reference to India and other developing countries | treatment of participants with COVID19 | | clinical studies on COVID-19<br><br>Narrative review |
| Kapoor et al., 2020 | 30-March-2020 | Role of Chloroquine and Hydroxychloroquine in the Treatment of COVID-19 Infection- A Systematic Literature Review | To summarize the available evidence regarding the role of chloroquine in treating coronavirus infection. | Systematic review (SR) | Did not include completed clinical studies on COVID-19 |
| Gbinigie et al., 2020 | 7-April-2020 | Should chloroquine and hydroxychloroquine be used to treat COVID-19? A rapid review | To establish the current evidence for the effectiveness of CQ and HCQ in treating COVID-19 infection | Narrative review | Narrative review<br><br>Did not include completed clinical studies on COVID-19 |
| Pastick et al, 2020 | 13-April-2020 | Review: Hydroxychloroquine and Chloroquine for Treatment of SARS-CoV-2 (COVID-19) | A review of all the available evidence of safety and efficacy of HCQ & CQ | Narrative review | Narrative review |
| Shukla et al., 2020 | 28-April-2020 | Chloroquine and hydroxychloroquine in the context of COVID-19 | To present the available in vitro and clinical data for the role of chloroquine/ hydroxychloroquine in COVID-19 and attempts to put them into perspective, especially in relation to the different risks/benefits particular to each patient who may require treatment. | Narrative review | Narrative review |
| Hashem et al., 2020 | 29-April-2020 | Therapeutic use of chloroquine and hydroxychloroquine in COVID-19 and other viral infections: A narrative review | To comprehensively review previous studies which used CQ or HCQ as an antiviral treatment. | Narrative review | Narrative review |
| Patil et al., 2020 | 11-May-2020 | A systematic review on use of aminoquinoline for the therapeutic management of COVID-19: Efficacy, safety and clinical trials | Provides a systematic review of mechanism of action, efficacy, and safety of chloroquine and hydroxychloroquine which are being used as therapeutic measure to cure COVID-19 infection. | Systematic review (SR) | Did not include completed clinical studies on COVID-19 |

**Supplementary Table S3. Quality Assessment for Included Reviews**

| | Methodological Quality Assessment of the included studies Systematic Reviews - AMSTAR Items | | | | | | | | | | | |
|---|---|---|---|---|---|---|---|---|---|---|---|---|
| | **1** | **2** | **3** | **4** | **5** | **6** | **7** | **8** | **9** | **10** | **11** | **Rating** |
| Takla 2020 | 1 | 1 | 1 | 1 | 1 | 1 | 1 | 1 | 0 | 0 | 1 | 9 |
| Yang 2020 | 1 | 1 | 1 | 1 | 0 | 1 | 1 | 1 | 1 | 1 | 1 | 10 |
| Jankelson 2020 | 1 | 1 | 1 | 1 | 0 | 1 | 0 | 0 | 0 | 0 | 0 | 5 |
| Shamshirian | 1 | 0 | 1 | 1 | 1 | 1 | 1 | 1 | 1 | 1 | 1 | 10 |
| Sarma 2020 | 1 | 1 | 1 | 1 | 0 | 0 | 1 | 1 | 1 | 1 | 1 | 9 |
| Singh 2020 | 1 | 1 | 1 | 1 | 1 | 1 | 1 | 1 | 1 | 1 | 1 | 11 |
| Suranagi 2020 | 1 | 1 | 1 | 1 | 0 | 1 | 1 | 1 | 0 | 0 | 0 | 7 |
| Das 2020 | 1 | 0 | 1 | 1 | 1 | 1 | 1 | 1 | 0 | 0 | 1 | 9 |
| Chowdhury 2020 | 1 | 1 | 1 | 1 | 1 | 1 | 1 | 1 | 0 | 0 | 1 | 9 |
| Chacko 2020 | 1 | 1 | 1 | 1 | 1 | 1 | 1 | 1 | 1 | 0 | 1 | 10 |
| Hernandez 2020 | 1 | 1 | 1 | 1 | 1 | 1 | 1 | 1 | 0 | 0 | 0 | 8 |
| Rodrigo 2020 | 1 | 1 | 1 | 1 | 0 | 0 | 1 | 1 | 0 | 0 | 1 | 7 |
| Wang 2020 | 1 | 1 | 0 | 1 | 1 | 1 | 0 | 0 | 0 | 0 | 1 | 6 |

**Supplementary Table S4: Characteristics of Excluded Primary Studies**

| Study and date of publication | Country | Reviews | Reason for exclusion |
|---|---|---|---|
| Bessière et al., (1-May-2020) | France | 8 | Observational studies |
| Borba et al., (24-April-2020) | Brazil | 6, 8, 11, 12 | Comparison between two different doses of CQ (not SOC) |
| Carlucci et al., (8-May-2020) | USA | 9 | Observational study |
| Chorin et al., (1-May-2020) | USA and Italy | 8 | Observational study |
| Chorin et al., (3-April-2020) | USA | 2, 4, 8, 9, 11, 12, | Observational study |
| Gao et al., (16-March-2020) | China | 7, 13 | No data presented |
| Gautret et al., (11-April-2020) | France | 1, 2, 3, 4, 8, 9, 10, 12, 13 | Observational study |
| Geleris et al., (7-May-2020) | USA | 4, 7, 8, 9, 10, 11 | Observational study |
| Huang et al., (4-May-2020) | China | 8 | Observational study |
| Ip et al., (25-May-2020) | USA | 11 | Observational study |
| Jiang et al., (12-July-2020)?? | China | 13 | No full text - abstract only |
| Kim et al., (18-May-2020) | South Korea | 9, 11 | Observational study |
| Lee et al., (8-May-2020) | South Korea | 9 | Observational study |
| Magnagnoli et al., (23-April-2020) | USA | 3, 4, 5, 7, 8, 9, 10, 11, | Observational study |
| Mallat et al., (2-May-2020) | UAE | 7, 8, 9, 11 | Observational study |
| Mehra et al., (22-May-2020) | USA | 9, 11 | Observational study which was retracted |
| Membrillo et al., (9-May-2020) | Spain | 8, 9 | Observational study |
| Mercuro et al., (1-May-2020) | USA | 8, 10, 11 | Observational study |
| Million et al., (5-May-2020) | France | 1, 3, 9, 10, 11, 12, 13 | Observational study |
| Molina et al., (17-April-2020) | France | 1, 3, 4, 8, 9, 10, 12, 13 | No control group |
| Okour et al., (13-May-2020) | USA | 9 | Duplicate data |
| Perinel et al., (7-April-2020) | France | 12 | Observational study |
| Ramireddy et al., (25-April-2020) | USA | 8, 9, 10, 11 | Case series |
| Raoult et al., (11-April-2020) | France | 8 | Case series |
| Regina et al., (12-May-2020) | Switzerland | 9 | Observational study |

| | | | |
|---|---|---|---|
| Rosenberg et al., (11-May-2020) | USA | 7, 9, 11 | Observational study |
| Saleh et al., (29-April-2020) | USA | 8, 9, 11 | Observational study |
| Singh et al., (19-May-2020) | USA | 9, 11 | Observational study |
| Van den Broek et al., (29-April-2020) | Netherlands | 8 | Observational study |
| Yu et al., (1-May-2020) | China | 7, 8 | Observational study |
| Yu et al., (15-May-2020) | China | 11 | Observational study |
| Esper et al., | Brazil | None | COVID not confirmed by PCR |
| **Barbosa et al, (15-April-2020)** | Brazil | None | 1. Design not clear<br>2. Both groups of participants received HCQ |
| **Mahevas et al, (14-April-2020)** | France | None | Observational study |
| **Skipper et al, (16-July-2020)** | Canada | None | Covid-19 not confirmed with PCR |
| **Cavalcanti et al, 23-July-2020)** | Brazil | None | Some participants had suspected Covid-19 not confirmed by PCR |

**Supplementary Table S5. Assessment of risk of bias in included experimental studies**

| Safeguard item | S 1 | S 2 | S 3 | S 4 | S5 | S 6 | S 7 | S 8 |
|---|---|---|---|---|---|---|---|---|
| | Chen Jun et al. [illegible] | Chen Z et al. [illegible] | Gautret et al. [illegible] | Tang et al. 2020 | Huang et al 2020 | Horby et al. 2020 | Oriol et al. 2020 | Chen P.C. et al [illegible] |
| 1. Data collected after the start of the study was not used to exclude participants or to select them into the analysis | 1 | 1 | 1 | 1 | 1 | 1 | 1 | 1 |

| | | | | | | | | |
|---|---|---|---|---|---|---|---|---|
| 2. Participants in all comparison groups were from the same population and timeframe | 1 | 1 | 0 | 1 | 1 | 1 | 1 | 1 |
| 3. Inclusion/ exclusion criteria specified and applied equally to all groups prior to group assignment | 1 | 1 | 0 | 1 | 1 | 1 | 1 | 1 |
| 4. Any attrition (or exclusions after entry) is less than 20% of total participant numbers | 1 | 1 | 0 | 1 | 1 | 1 | 1 | 1 |
| 5. Missing data is less than 20% | 1 | 1 | 1 | 1 | 1 | 1 | 1 | 1 |
| 6. Analysis accounted for missing data | 1 | 1 | 0 | 1 | 1 | 1 | 1 | 1 |
| 7. Treatment deviations or non-compliance/ non-adherence were less than 20% | 1 | 1 | 1 | 1 | 1 | 1 | 1 | 1 |
| 8. Analysis accounted for treatment deviations/ withdrawals | 1 | 1 | 0 | 1 | 1 | 1 | 1 | 1 |
| 9. Procedures for data collection of covariates were reliable and the same for all participants | 1 | 1 | 0 | 1 | 1 | 1 | 1 | 1 |
| 10. Outcome was objectively defined | 1 | 1 | 1 | 1 | 1 | 1 | 1 | 1 |
| 11. Analyst was blinded | 0 | 1 | 0 | 1 | 1 | 1 | 0 | 0 |
| 12. Outcome assessor(s) were blinded | 0 | 1 | 0 | 0 | 0 | 0 | 1 | 0 |
| 13. Participants were blinded | 0 | 1 | 0 | 0 | 0 | 0 | 0 | 0 |
| 14. Caregivers were blinded | 0 | 1 | 0 | 0 | 0 | 1 | 0 | 0 |
| 15. Exposures/ interventions were objectively defined | 1 | 1 | 1 | 1 | 1 | 1 | 1 | 1 |
| 16. Care was delivered equally to all participants | 1 | 1 | 0 | 1 | 1 | 1 | 1 | 1 |
| 17. Cointerventions that could impact the outcome were comparable between groups or avoided | 1 | 1 | 0 | 1 | 1 | 1 | 1 | 1 |
| 18. Control and active interventions/ exposures are sufficiently distinct | 1 | 1 | 1 | 1 | 1 | 1 | 1 | 1 |
| 19. Exposure/intervention definition consistently applied to all participants | 1 | 1 | 1 | 1 | 1 | 1 | 1 | 1 |
| 20. Outcome definition consistently applied to all participants | 1 | 1 | 1 | 1 | 1 | 1 | 1 | 1 |
| 21. The time period between exposure and outcome is similar across patients and between groups or the analyses adjust for different lengths of follow-up of patients | 1 | 1 | 1 | 1 | 1 | 1 | 1 | 1 |
| 22. Design features in place that account for confounding | 1 | 1 | 0 | 1 | 1 | 1 | 1 | 1 |
| 23. Analytic strategies in place for key confounders | 0 | 0 | 0 | 1 | 0.5 | 1 | 0 | 1 |
| 24. Key baseline characteristics / prognostic indicators for the study were comparable across groups | 1 | 1 | 0 | 1 | 0.5 | 1 | 1 | 1 |
| 25. Allocation procedure was adequately concealed | 1 | 0 | 0 | 0 | 0 | 1 | 1 | 0 |

| | | | | | | | | |
|---|---|---|---|---|---|---|---|---|
| 26. Conflict of interests were declared and absent | 0 | 1 | 1 | 1 | 1 | 1 | 1 | 1 |
| 27. Participants were randomly allocated to groups with adequate randomisation process | 1 | 1 | 0 | 1 | 0.5 | 1 | 1 | 1 |
| 28. Analytic method was justified by study design | 1 | 1 | 0 | 1 | 0.5 | 1 | 1 | 1 |
| 29. Computation errors or contradictions were absent | 1 | 1 | 1 | 1 | 1 | 1 | 1 | 1 |
| 30. There was no data dredging or selective reporting of the outcome | 1 | 1 | 1 | 1 | 1 | 1 | 1 | 1 |
| 31. All subjects were selected prior to intervention/exposure and evaluated prospectively | 1 | 1 | 0 | 1 | 1 | 1 | 1 | 1 |
| 32. Carry-over or refractory effects were avoided or considered in the design of the study or were not relevant | 1 | 1 | 1 | 1 | 1 | 1 | 1 | 1 |
| 33. The intervention/ exposure period was long enough to have influenced the study outcome | 1 | 1 | 1 | 1 | 1 | 1 | 1 | 1 |
| 34. Dose of intervention/ exposure was sufficient to influence the outcome | 1 | 1 | 1 | 1 | 1 | 1 | 1 | 1 |
| 35. Length of follow-up was not too long or too short in relation to the outcome assessment | 1 | 1 | 1 | 1 | 1 | 1 | 1 | 1 |
| Summary count of safeguard items | 29 | 33 | 16 | 31 | 29 | 33 | 31 | 30 |

#### Supplementary Fig. 1: Quality Assessment

**A: AMSTAR Assessment for Included Reviews**

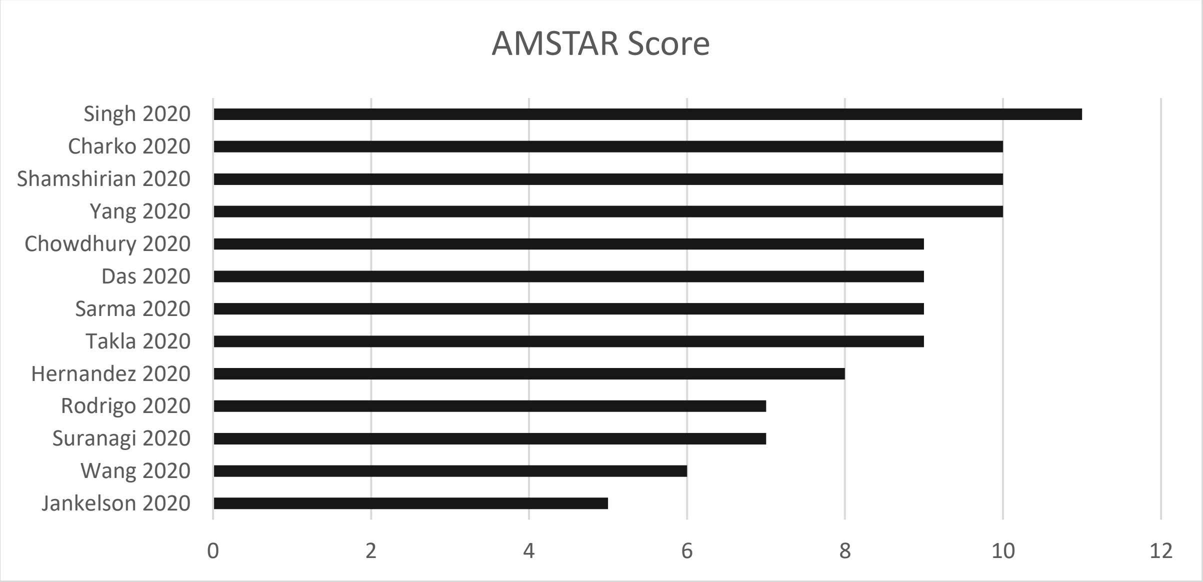

**B: MASTER for experimental studies**

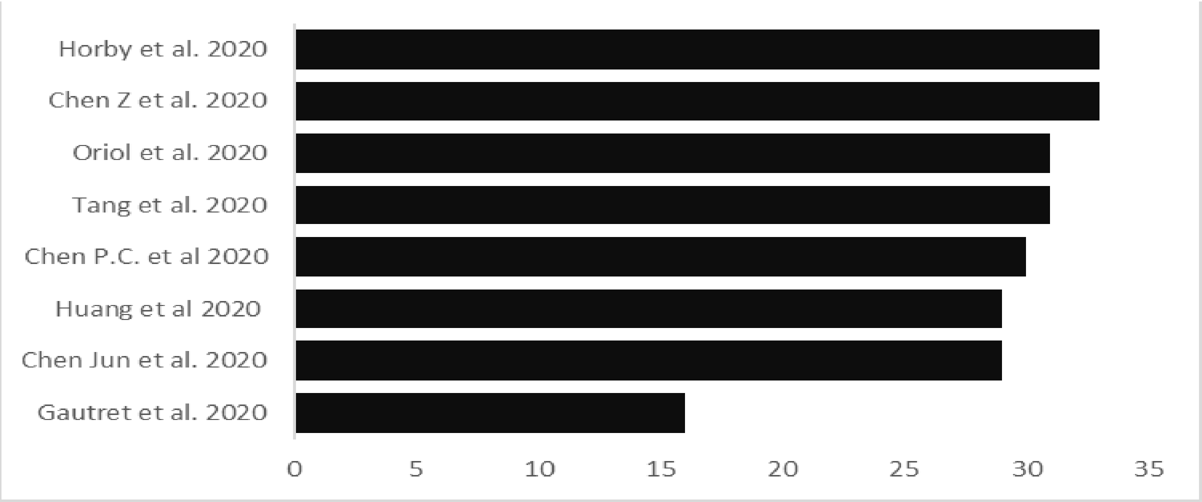

#### Supplementary Table S6. Outcomes of Included reviews

| Author, date & design | Drug | Mortality | ICU, intubation | Virological Cure | Disease worsening | Adverse events |
|---|---|---|---|---|---|---|
| Shamshirian (28 May 2020)<br><br>SRMA | HCQ | No significant difference in mortality (RR: 1.13, 95%CI 0.71 - 1.80) | Intubation - no significant differences HCQ - OR: 2.11, 95% CI, 0.31-14.03, $I^2$ =75.6%) | HCQ - No effectiveness (RR: 0.96, 95% CI, 0.76-1.22), (RD: 0.00, 95% CI, - 0.18-0.18) | No considerable disease exacerbation difference HCQ (RR: 0.59, 95% CI, 0.04-7.79) | Higher risk of events HCQ (OR: 4.01, 95% CI, 1.17-13.84 |
| | HCQ+AZI | Higher mortality in HCQ+AZI (RR 2.46, 95% CI 1.4-4.3) | - | HCQ +AZI - No significant difference with control (RR 2.15, 95%CI 0.31 - 14.77) | | |
| Chacko (20 May 2020)<br><br>SRMA | HCQ | No significant difference in mortality (OR: 1.41, 95% CI: 0.76 – 2.62) | - | No significant different between the HCQ and control groups (OR: 1.13, CI: 0.26–5.01; p = 0.87) | No difference in clinical worsening (OR 1.1, 95% CI: 0.6–2.02). | Higher risk of adverse events in HCQ arm (OR: 4.1, CI: 1.42 – 11.88; p = 0.009) |
| Yang (14 May 2020)<br><br>SRMA | HCQ+AZI | Increased mortality in HCQ +/- AZI (OR 2.5 (95% CI 1.4 - 4.5) | - | No difference between HCQ alone vs. control (OR = 1.74 95% CI 0.51 - 5.91) | No significant difference between HCQ alone versus Control (OR = 1.37, 95% CI 0.09 - 21.97) | |
| | HCQ | HCQ alone was associated with increased mortality (OR 2.98 (95%CI 1.6 – 5.7) | - | No difference between HCQ with or without AZI vs control (OR = 1.95, 95% CI 0.19 - 19.73) | No difference between HCQ+AZI versus Control (OR = 1.00, 95% CI 0.27 – 3.75) | |
| Singh (7 May 2020)<br><br>SRMA | HCQ | Higher mortality in HCQ arm (RR, 2.17; 95% 1.32 to 3.57) | No difference in transfer to the ICU (20.2 vs 22.1%; RR 0.91, 0.47–1.80) | No benefit with HCQ (RR, 1.05; 95% CI, 0.79 to 1.38; p=0.74) | Improvement in pneumonia with HCQ (80.6 vs 54.8%, p=0.048), chest CT with HCQ (61.3 vs 16.1%) | 1 study reported no adverse events. 1 reported no cardiac toxicity although it did not report how they assessed this. 7 other studies reported moderate to large increases in occurrence of adverse events |

| | | | | | | |
|---|---|---|---|---|---|---|
| Sarma (13 April 2020)<br><br>SRMA | HCQ | No difference in death or clinical worsening between treatment and control group (OR, 1.37, 95% CI, 0.09-21.97) | - | No difference in virological cure 2.37; 95% CI, [0.13-44.53] | 2 studies found shorter time for body temperature normalization and the number of cough days. 3 studies reported no difference in clinical worsening OR, 1.37 (95% CI, 0.09-21.97). | Reported adverse events but No significant difference on all studies (OR, 2.19; 95% CI, [0.59-8.18]) |
| | HCQ+AZI | | - | Higher virological cure in 1 study while others reported moderate effect of the combination | - | Reported adverse events and QTc prolongation. 1 study found no signs of cardiac toxicity |
| Wang (1 June 2020)<br><br>SR | HCQ-AZI | One study had no deaths, second study had lower mortality in HCQ-AZ group, but was statistically non-significant. | Marginally lower percentage in HCQ were transferred to ICU but differences were not significant | - | HCQ improved pneumonia: 80.6% vs 54.8%; (1 study), CQ reduced exacerbation of pneumonia, improved lung imaging, promoted virological clearance, and shortened disease course. Poor clinical outcome significantly associated with greater severity (OR 10.05) in one study. | More adverse events in HCQ and CQ than control like QTc interval prolongation. 2 studies reported no serious adverse events |
| Takla (30 May 2020)<br><br>SR | CQ/HCQ | 60% of studies reported no difference on mortality, 30% reported higher mortality in the HCQ group, 10% reported reduced mortality in HCQ group | No difference in need for mechanical ventilation, and transfer to an intensive care unit | 67% of studies showed significant higher viral clearance, 33% no difference | - | A higher probability of adverse events in 82% of the studies |
| Das (28 May 2020)<br><br>SR | HCQ | Reported no significant effect of HCQ on death | Reported no significant effect of HCQ on intubation | 5 studies (1269 participants) reported good virological and clinical outcomes in the HCQ arm<br>5 studies (808 participants) showed negative or equivocal results | Reported "good virological and clinical outcomes in the HCQ arm" | 4 studies (1207 participants) reported HCQ as safe with mild adverse events.<br>2 studies (101 participants) reported QT prolongation associated with HCQ treatment<br>5 studies (859 participants) reported HCQ associated with serious adverse events |

| | | | | | | |
|---|---|---|---|---|---|---|
| Hernandez (27 May 2020)<br><br>SR | HCQ<br>CQ | 1 study reported no deaths, 2 studies found decreases in mortality, 2 found no change in mortality, 4 found moderate to large increases in mortality | 2 studies reported CQ/HCQ had increased need for ICU admission, intubation and/ mechanical ventilation.<br>3 studies reported no effect of CQ/HCQ on ICU, intubation or need for mechanical ventilation | 2 studies reported moderate to large increases in virologic clearance for the CQ/HCQ arm. 3 studies found no difference or effect. 1 study found large decreases of virologic clearance in the CQ/HCQ arm | 1 study reported increased progression of the disease (progressing to need respiratory support), 1 study reported fewer participants in HCQ had disease progression and others had 1.0- and 1.1-day reduction in fever and cough. 3 studies found no effect. | 6 studies reported modest to large increase in adverse events and QTc prolongation in HCQ alone and HCQ+AZI group. 1 study reported both in adverse events and 1 study reported no difference and insufficient evidence of risk of adverse events |
| Rodrigo (16 May 2020)<br><br>SR | CQ<br>HCQ | Mortality higher in higher dose of CQ arm (15% vs 39%, p=0.03) | - | No statistically significant difference by day 14 of illness (10/10 in CQ group vs. 11/12 in Lopinavir). The other study terminated early | - | - |
| | | | | 3 studies reported no statistical difference in clearance of viremia<br>1 study reported statistical significance (70% in HCQ group vs. 12.5% in placebo group, p=0.001) | - | - |
| Suranagi (13 May 2020)<br><br>SR | HCQ | HCQ increased risk of death | 1 study reported intubations, but No significant effect of HCQ on risk of either mechanical ventilation or intubation | 3 studies reported HCQ reduced viral load and effect reinforced by Azithromycin. 1 study reported no effect on viral clearance | HCQ reduced duration of illness and improved pneumonia and pulmonary image changes | - |
| Chowdhury (28 April 2020)<br><br>SR | CQ<br>HCQ | - | - | Better virological clearance in in the CQ/HCQ arm than control (standard care/Lopinavir/ritonavir arm). | Improved pneumonia per chest CT and reduced progression to severe illness. 2 studies reported no significant different in alleviating or increasing severity of disease | Adverse effects CQ arm but not in Lopinavir/ritonavir arm, (n= 1 study), HCQ arm (Adverse events: 30% vs 8.8 %). 1 study found some minor adverse events in HCQ arm. 1 study sound no significant difference on adverse events between groups. |
| Jankelson (31 May 2020)<br><br>SR | CQ/HCQ | - | - | - | - | QTc prolongation in 40 participants<br><br>Ventricular arrythmia reported in 2 participants, first degree AV |

| | | | | | | |
|---|---|---|---|---|---|---|
| | | | | | | block developed in 1 and LBBB in another patient |

#### Supplementary Table S7. Limitations and conclusions from reviews

| Author name & date | Review Conclusion | Limitations of the review |
|---|---|---|
| Shamshirian (28 May 2020) | No clinical benefits regarding HCQ treatment with/without azithromycin for COVID-19 participants | Few included studies (6 studies) with small sample sizes in the meta-analysis |
| Chacko (20 May 2020) | Meta-analysis does not support the treatment of COVID-19 infection with HCQ | Small studies, small sample sizes, different outcome measurements and endpoints measured at different intervals |
| Yang (14 May 2020) | HCQ with or without AZI are beneficial for treatment of COVID-19 participants, but may also have higher mortality | Few included studies with small sample sizes. |
| Singh (7 May 2020) | While no benefit on viral clearance demonstrated by HCQ, a significant 2-fold increase in mortality with the HCQ warrants its use, if at all, with extreme caution. | Small number of participants overall, combining the results of RCT with other non-randomized studies. |
| Sarma (13 April 2020) | Treatment with HCQ may result in reducing radiological progression with comparable safety | Few studies, small sample sizes, and several studies without controls |
| Wang (1 June 2020) | No solid evidence supporting the efficacy and safety of HCQ and CQ as a treatment for COVID-19 with or without azithromycin | Search not comprehensive, only searched 2 English databases, few studies with small sample sizes |
| Jankelson (31 May 2020) | Compelling evidence that CQ & HCQ induce significant QTc prolongation and can be potential risk factors of arrythmia. | Small number of studies, small sample sizes. Efficacy outcomes not part of scope of review |
| Takla (30 May 2020) | Relative to standard in-hospital management of symptoms, the use of CQ and HCQ to treat hospitalised COVID-19 has likely been unsafe | Data synthesize by describing percentage of studies with outcome not optimal |
| Das (28 May 2020) | Inconclusive evidence of HCQ efficacy and safety | Lack of data from well-designed RCTs |
| Hernandez (27 May 2020) | Insufficient and often conflicting evidence on the benefits and harms of using hydroxychloroquine or chloroquine to treat COVID-19. | There were few controlled studies, and control for confounding was inadequate in observational studies. |
| Rodrigo (16 May 2020) | Role of CQ & HCQ in covid19 is yet unclear and needs to be assessed by well-designed double-blind clinical trails | Small number of studies that were not similar hence a meta-analysis was not performed |
| Suranagi (13 May 2020) | Current evidence stands inadequate to support the use of hydroxychloroquine in pharmacotherapy of COVID-19 | Small studies with weak study designs |
| Chowdhury (28 April 2020) | No enough data to support the routine use of either HCQ or CQ for treatment of COVID19 | Narrative review. Indiscriminate inclusion criteria |

#### Random Effects Models

##### Mortality – random effects model

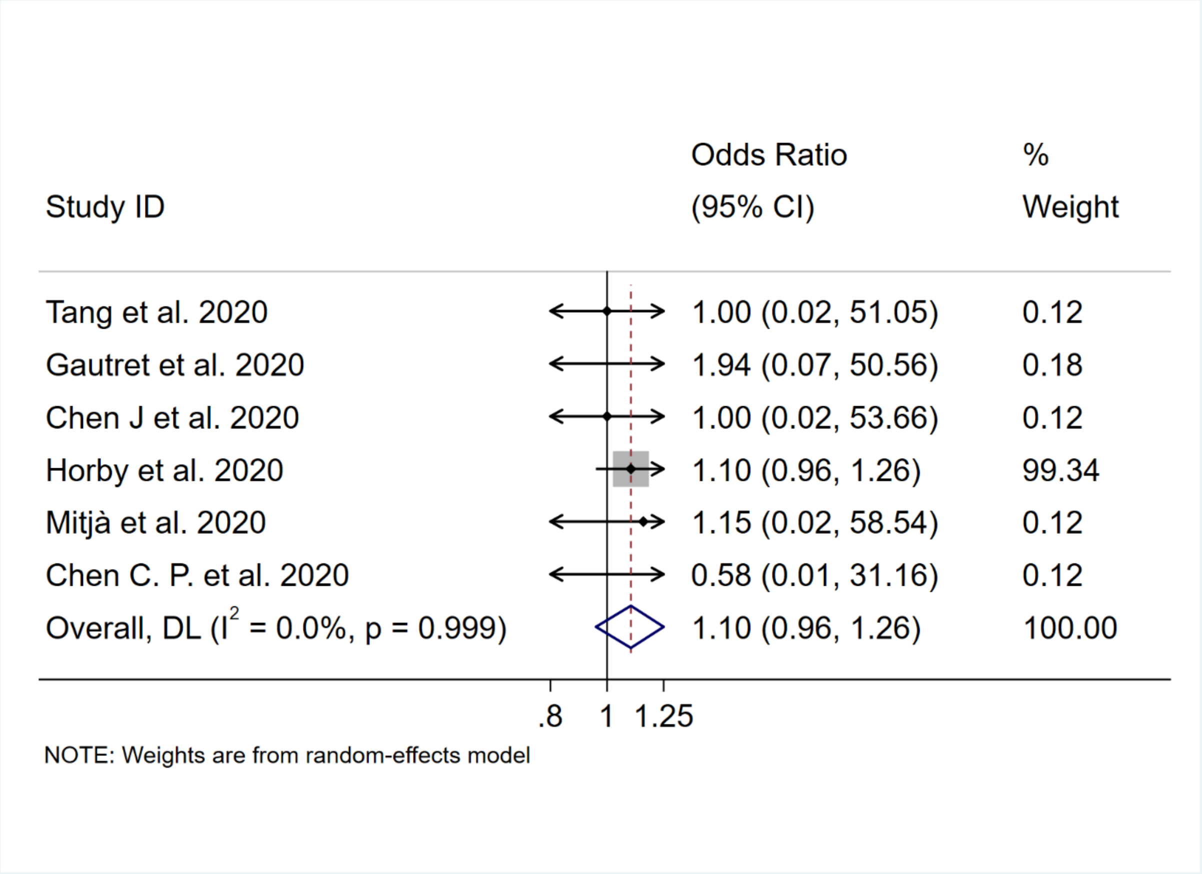

#### Mortality – excluding Horby et al. (2020)

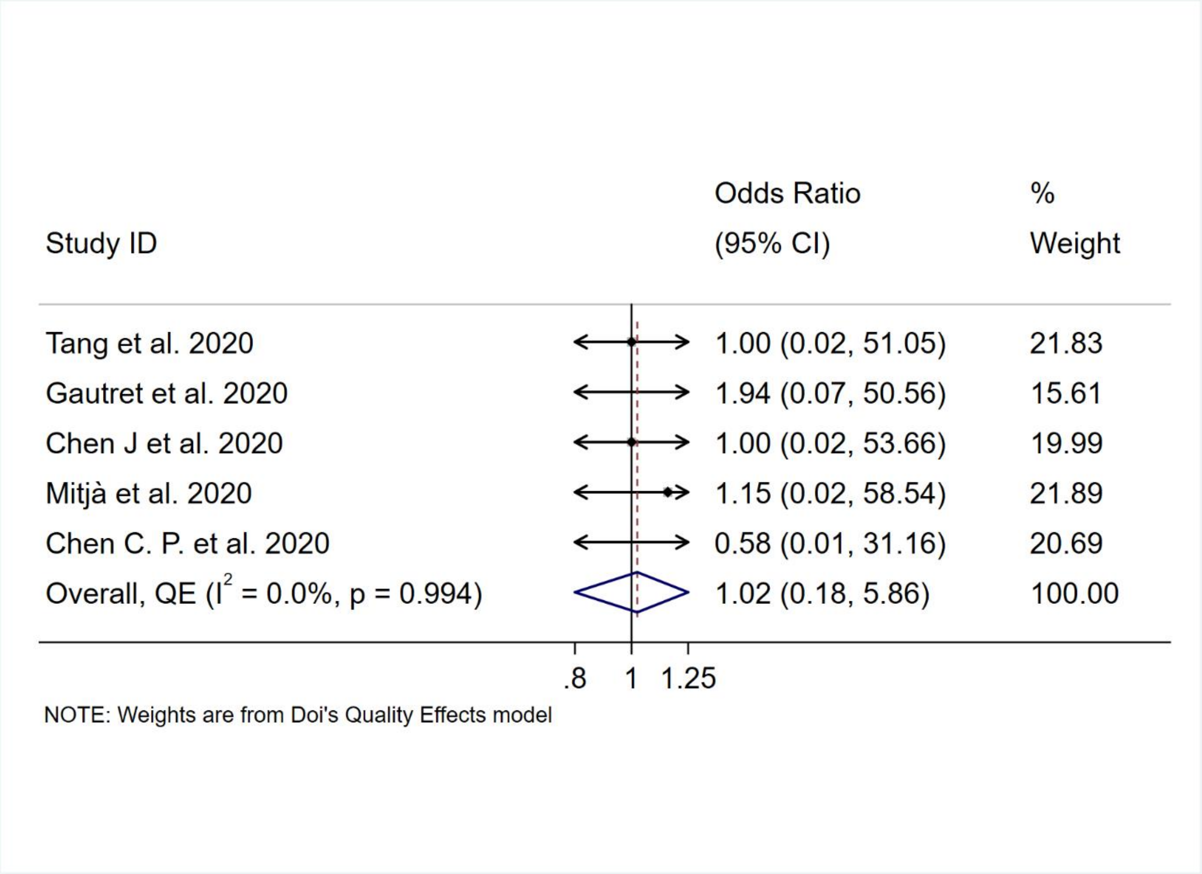

#### ICU – random effects model

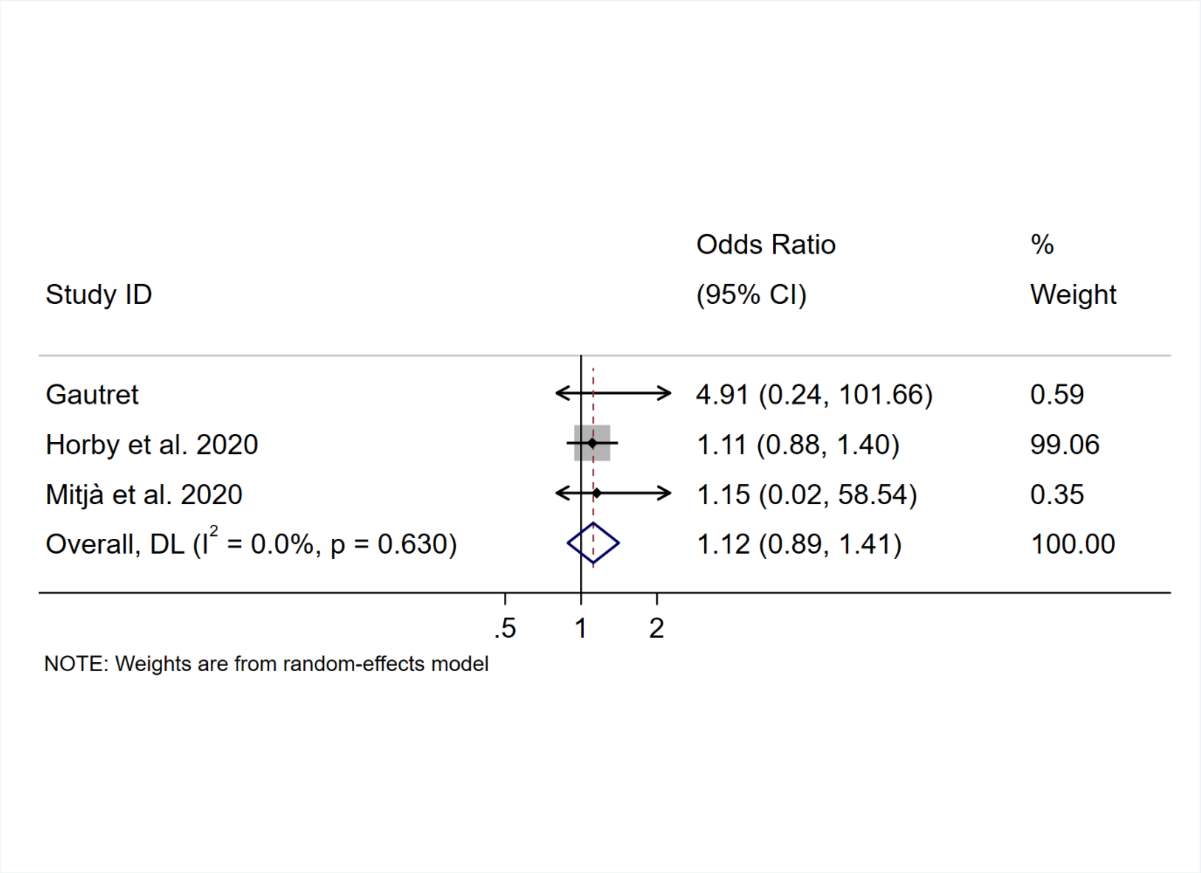

#### Virological cure – random effects

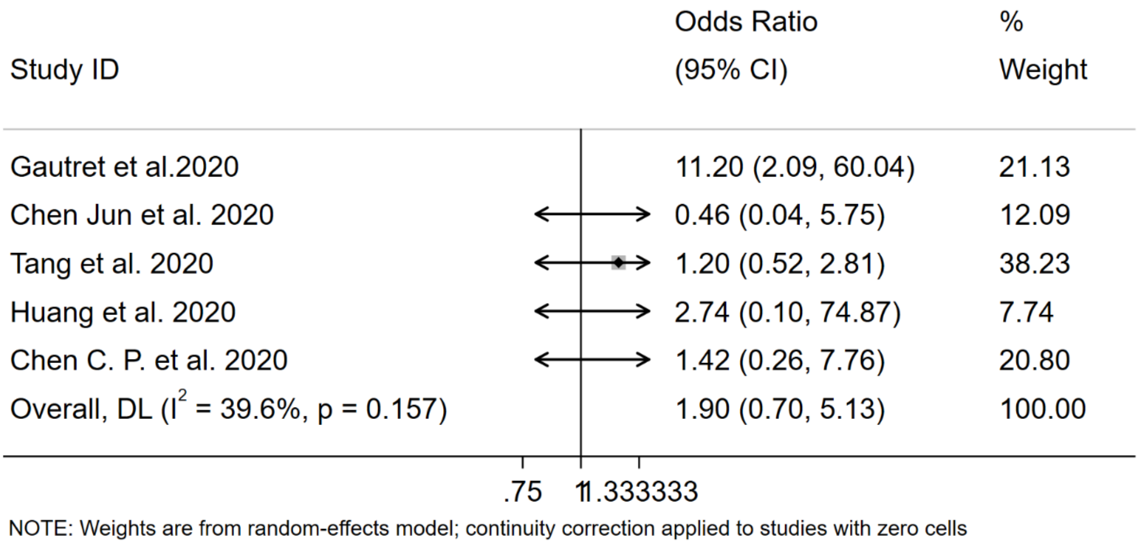

#### Virological cure – without Gautret

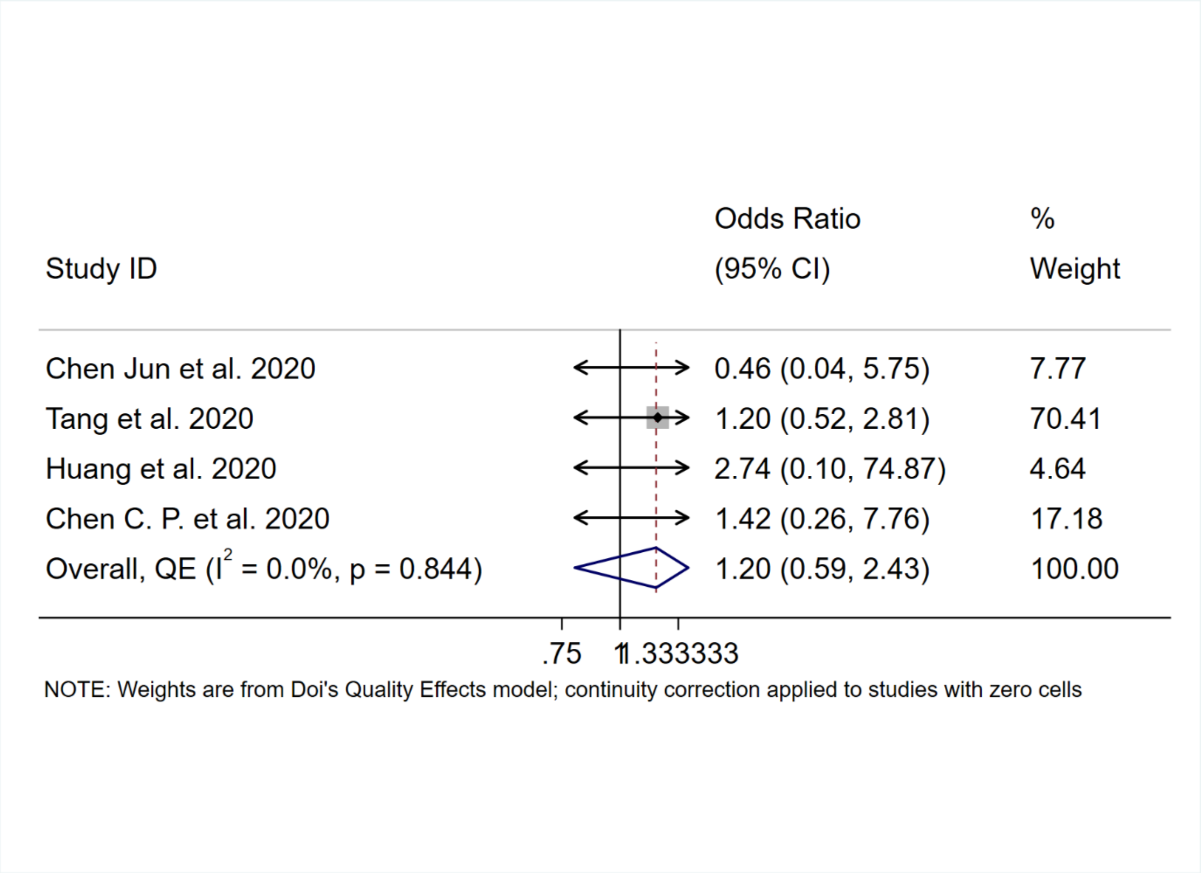

#### Disease worsening - random effects model

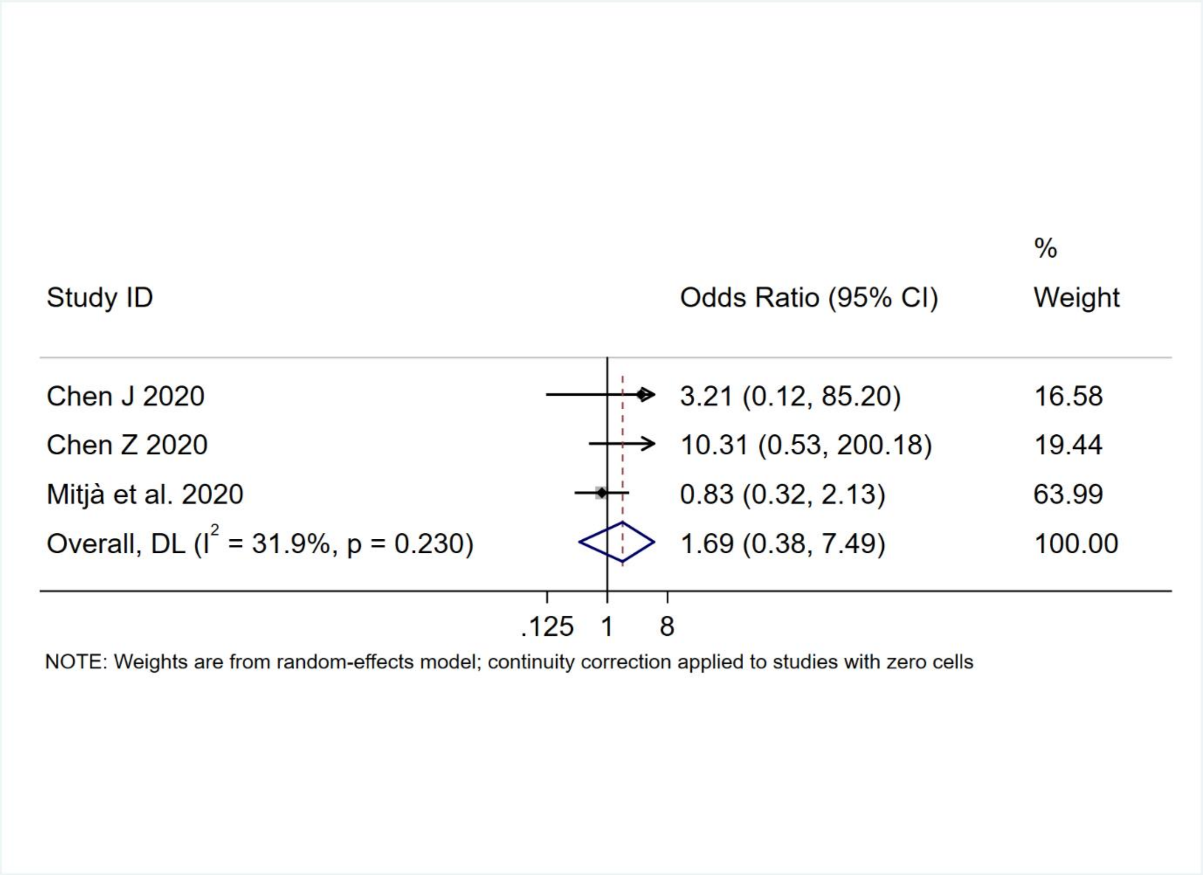

Adverse events – random effects

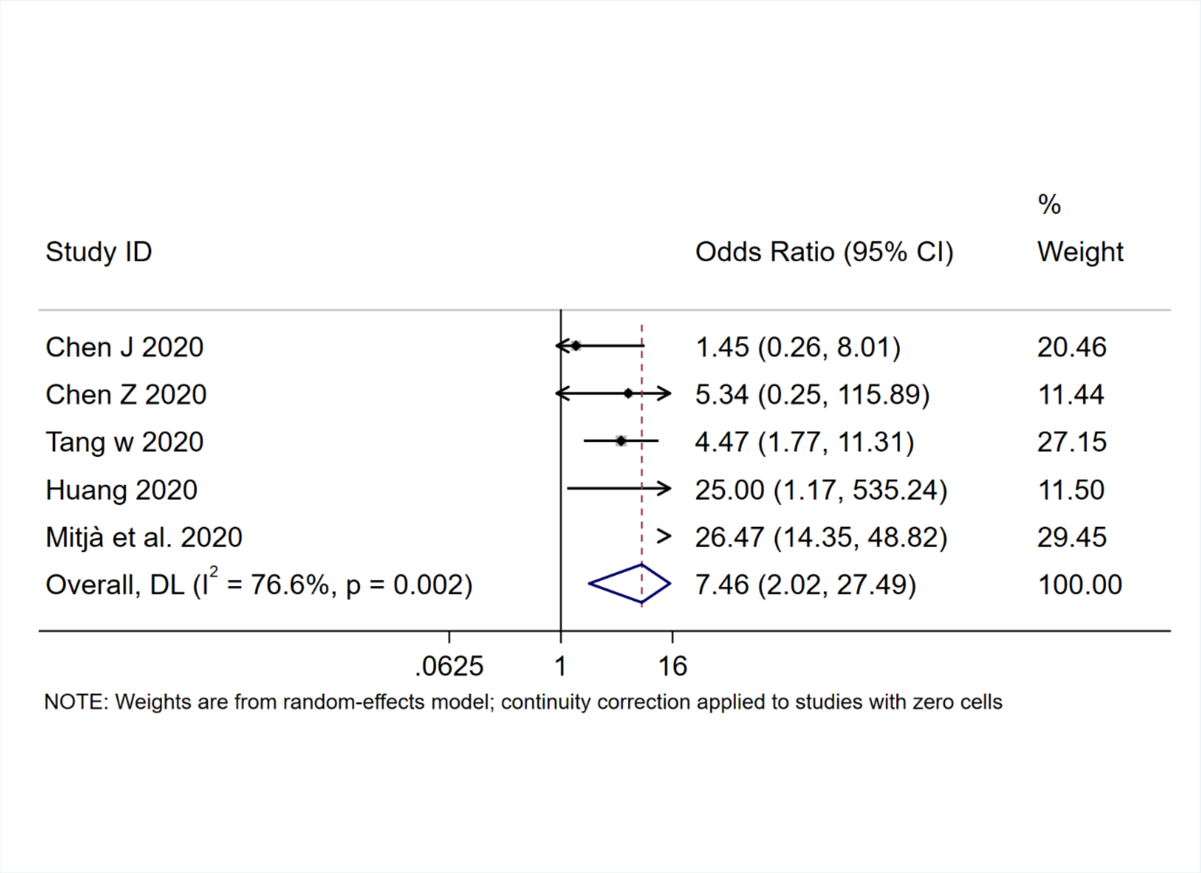

**Supplementary Fig. 3: Doi Plots and LFK Index**

3A: Mortality

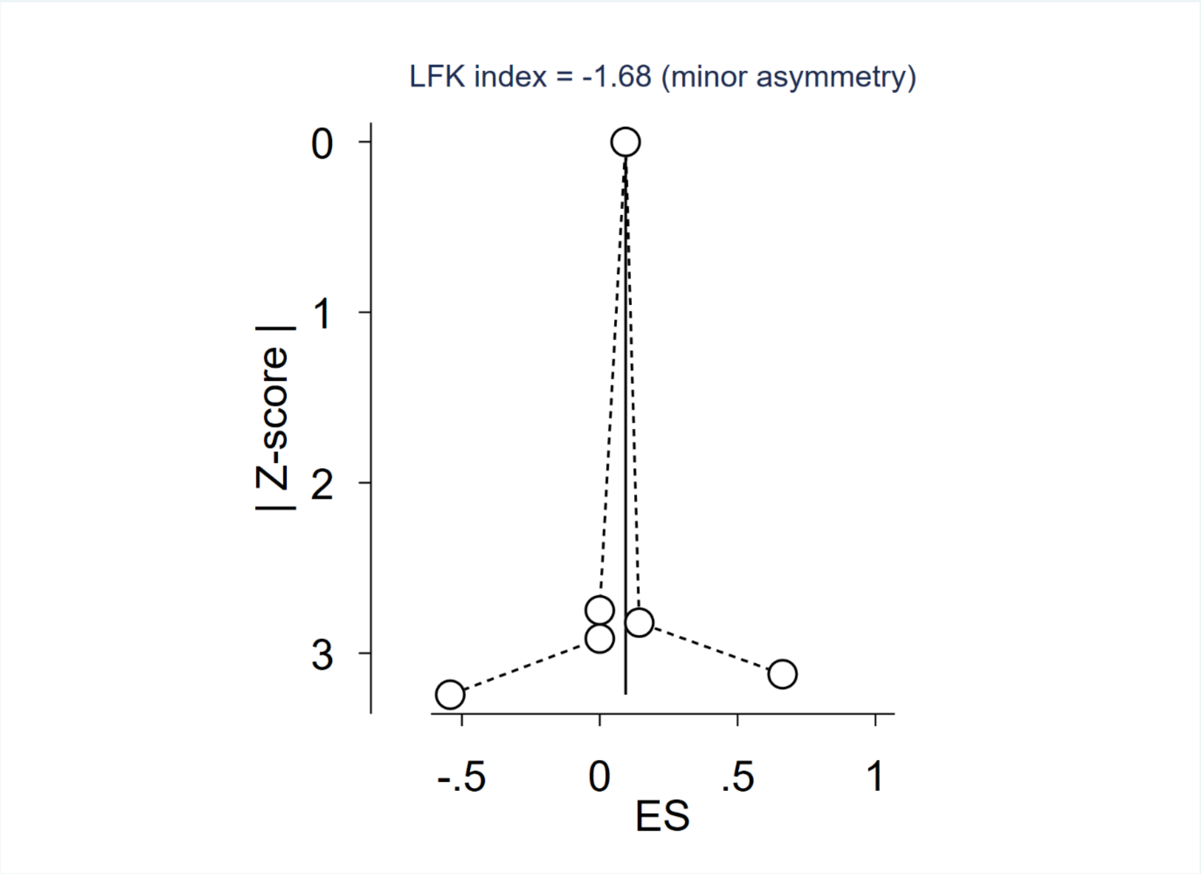

3B: ICU Admission

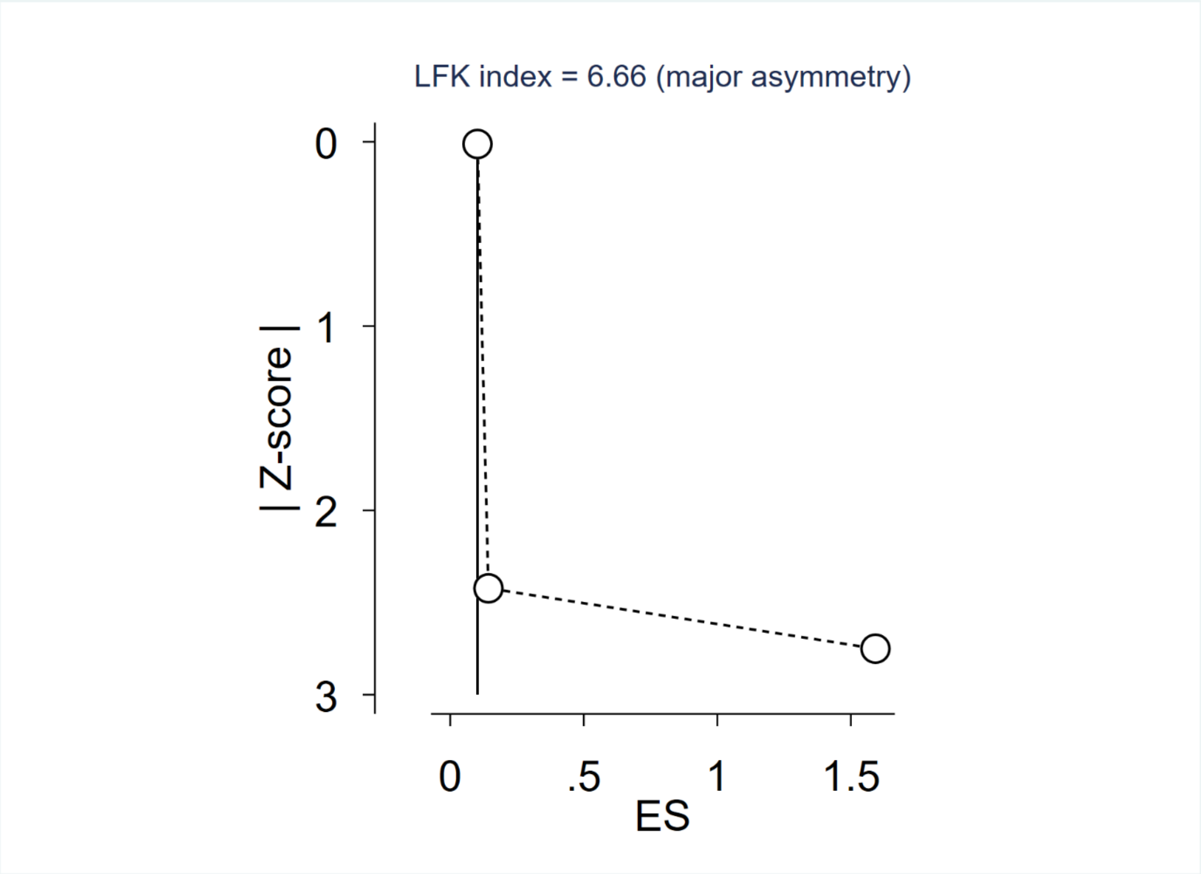

3C: Virological cure

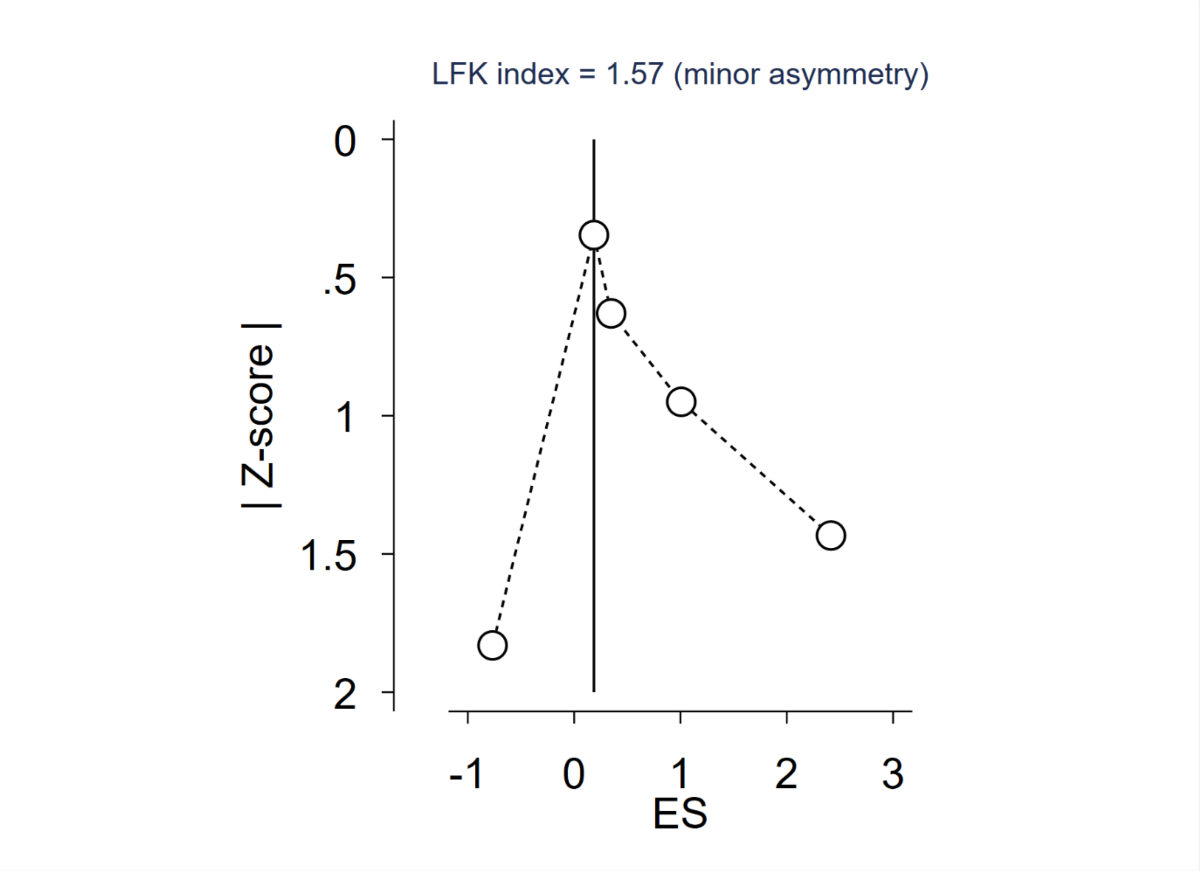

3D: Disease Exacerbation

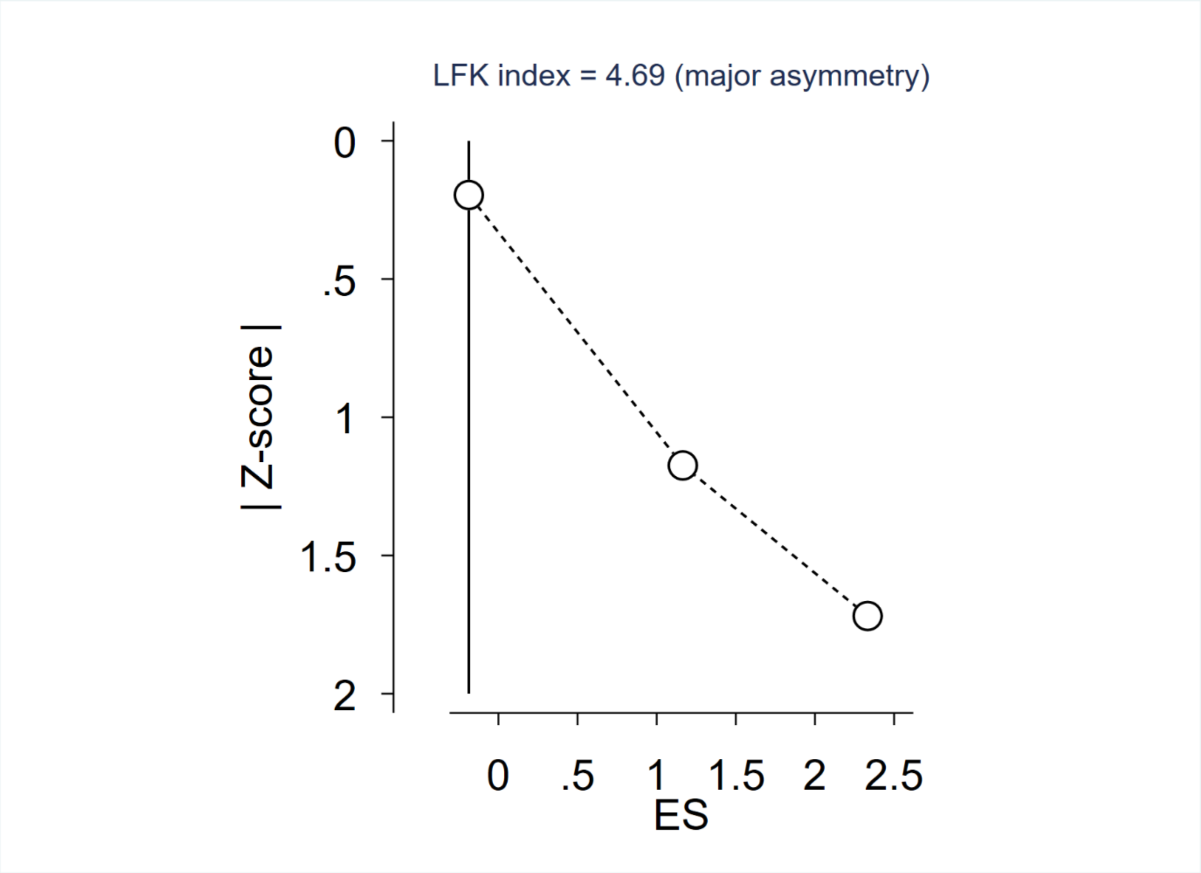

3E: Adverse events

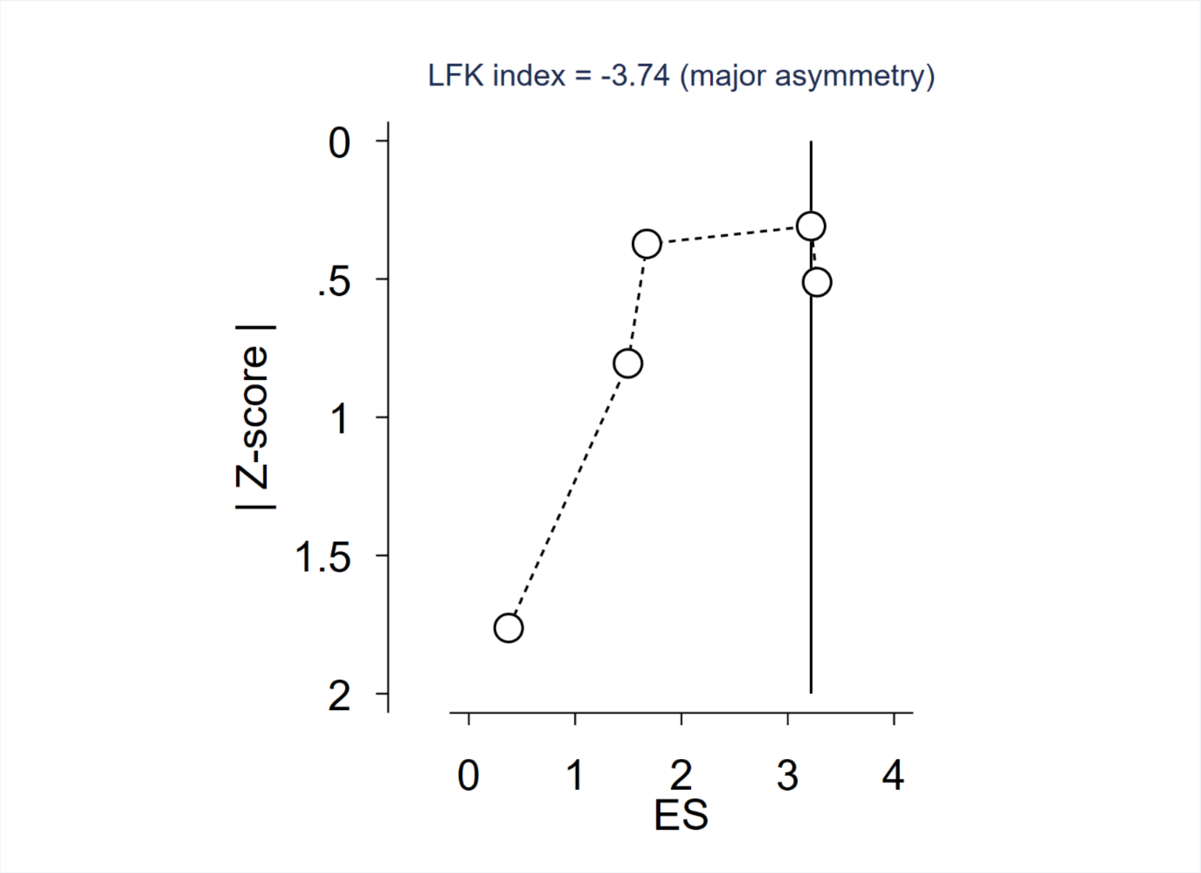
